# Workplace hazards and health among informally employed domestic workers in 14 cities, United States, 2011-2012: using four approaches to characterize workers’ patterns of exposures

**DOI:** 10.1101/2022.06.27.22275951

**Authors:** Emily Wright, Jarvis T. Chen, Jason Beckfield, Nik Theodore, Nancy Krieger

## Abstract

**Objectives:** We characterized informally employed US domestic workers’ (DWers) exposure to patterns of workplace hazards, as well as singular hazards, and examined associations with DWers’ work-related and general health.

**Methods:** We analyzed cross-sectional data from the sole nationwide survey of informally employed US DWers with work-related hazards data, conducted in 14 cities (2011-2012; N=2,086). We characterized DWers’ exposures using four approaches: single exposures (n=19 hazards), composite exposure to hazards selected *a priori*, classification trees, and latent class analysis. We used city fixed effects regression to estimate the risk ratio (RR) of work-related back injury, work-related illness, and fair-to-poor self-rated health associated with exposure as defined by each approach.

**Results:** Across all four approaches—net of individual, household, and occupational characteristics and city fixed effects—exposure to workplace hazards was associated with increased risk of the three health outcomes. For work-related back injury, the estimated RR associated with heavy lifting (the single hazard with the largest RR), exposure to all three hazards selected *a priori* (did heavy lifting, climbed to clean, worked long hours) versus none, exposure to the two hazards identified by classification trees (heavy lifting, verbally abused) versus “No heavy lifting,” and membership in the most-versus least-exposed latent class were, respectively, 3.4 (95% confidence interval [CI] 2.7 to 4.1); 6.5 (95% CI 4.8 to 8.7); 4.4 (95% CI 3.6 to 5.3), and 6.6 (95% CI 4.6 to 9.4).

**Conclusions:** Measures of joint work-related exposures were more strongly associated than single exposures with informally employed US DWers’ health profiles.

**KEY MESSAGES:** *What is already known on this topic:* Informally employed domestic workers in the US and internationally are frequently exposed to physical and social hazards at work, but only two studies have quantitatively assessed these workers’ exposures to joint patterns of hazards, and neither examined such patterns in relation to health. What this study adds
We characterized informally employed US domestic workers’ exposure to 19 single hazards and to combinations of these hazards, using three distinct approaches: composite exposure to hazards selected *a priori*, classification trees, and latent class analysis. Across all approaches to defining exposure, domestic workers exposed to worse joint patterns of workplace hazards, as well as to certain single hazards, experienced greater risk of work-related back injury, work-related illness, and fair-to-poor self-rated health. How this study might affect research, practice, or policy
Results underscore the importance of conceptualizing and operationalizing measures that capture domestic workers’ patterns of exposures. Moreover, results support the use of a latent class approach for identifying potential subgroups of workers unduly burdened and—across multiple health metrics—harmed by employer practices.

## INTRODUCTION

In 2008, Krieger and colleagues formulated the “inverse hazard law,” stating that “the accumulation of health hazards tends to vary inversely with the power and resources of the populations affected.”[1] Developed in relation to their novel study of the combined health impacts of social and physical workplace hazards in a sample of low-wage US workers, this law highlights how exposures cluster together and jointly impact health, producing social inequities in health within designated occupational categories as well as occupational health inequities.[2-3] The inverse hazard law, thus, adds impetus for research examining the health impacts of— and comparing approaches for characterizing—the joint patterns of exposure experienced by workers.

Despite this, no quantitative studies in the US or internationally have gone beyond examining associations between exposure to single workplace hazards and health among informally employed domestic workers (DWers). As of 2019, at least 800,000 DWers were employed informally in housecleaning, childcare, or adult care work by private households, rather than by third-party agencies or government entities.[4-5] These DWers are subjected to numerous workplace hazards, including wage theft, verbal abuse, racial discrimination, heavy lifting, and more. Although these hazards are not unique to DWers, the overall frequency and social patterning of DWers’ exposures is likely distinct—reflecting their historical and contemporaneous exclusion from laws protecting workers’ rights and the wide range of employer practices and worker social characteristics within this occupational group.[6]

Research examining the health of DWers experiencing combinations of workplace hazards is urgently needed. The sparse literature quantitatively examining workplace exposures and health among DWers in the US and internationally has documented cross-sectional associations between various singular hazards (e.g., lack of rest breaks, cleaning chemicals, physical abuse, late wages) and work-related (e.g., back injury) and general health (e.g., self-rated health).[7-11] Our recent study using a novel latent class approach found that informally employed US DWers experience distinct patterns of workplace hazards, motivating further analyses of the health implications of such exposure patterns.[12] Finally, although a growing body of research indicates the conceptual and empirical importance of examining the health implications of workers’ joint exposures using a range of methods,[1, 13-14] few studies have employed multiple methods simultaneously.[15] Evidence regarding the utility of accessible composite exposure approaches and more complex model-based methods for characterizing workers’ joint exposures, in comparison to a single hazards approach, could inform future research on the health impacts of domestic work (DW) and related occupations. Moreover, evidence regarding which groups of DWers are potentially unduly harmed by employer practices could further inform ongoing work to enforce existing, and create new, DWer protections.

In this study, we used cross-sectional data from the sole nationwide survey of informally employed US DWers with work-related hazards data—conducted in 2011-2012 by the National Domestic Workers Alliance (NDWA), the University of Illinois Chicago Center for Urban Economic Development (UIC CUED), and the DataCenter—to characterize the health of DWers exposed to different combinations of workplace hazards. Informed by ecosocial theory and its emphasis on embodied integration of exposures,[3, 16, 17] we characterized DWers’ exposures using four approaches—single exposures, composite exposure to hazards selected *a priori*, classification trees, and latent class analysis—and used city fixed effects regression models to examine associations between exposure and work-related and general health.

## METHODS

### Study population

We analyzed cross-sectional data from the sole nationwide survey of informally employed US DWers with workplace hazards data (N=2,086), conducted from June 2011 to February 2012 in 14 cities (**Table 1**). NDWA, UIC CUED, and DataCenter investigators selected these cities to “represent every region of the country,”[18] ensure a large sampling frame of domestic workers, and make sure DWers’ community organizations in each city could serve as research partners. City-specific enrollment targets were derived from American Community Survey 2005-2009 five-year estimates (based on occupation, racialized group, and nativity)[19] and used to help ensure the sample reflected each city’s DWer labor force. The UIC Institutional Review Board approved this survey.

**Table 1.** **Observed characteristics, total and by domestic workers’ racialized group, citizenship and immigration status, and main occupation: National Domestic Workers Alliance and University of Illinois Chicago Center for Urban Economic Development data, 14 cities**,^**a**^ **United States, 2011-2012 (N = 2**,**086)**.

|  |  |  |  |  |  |  |  |  |  |  |  |
| --- | --- | --- | --- | --- | --- | --- | --- | --- | --- | --- | --- |
| Variable | Total | Racialized group |  |  |  | Citizenship and immigration status<sup>b</sup> |  |  | Main occupation |  |  |
|  |  | Latina/o | White | Black | "Asian or other" | US citizen | Doc. immigrant | Undoc. immigrant | House cleaning | Child care | Adult care |
| **Total number** | 2086 | 1168 | 553 | 187 | 178 | 761 | 537 | 644 | 1116 | 668 | 302 |
| **Individual characteristics** |  |  |  |  |  |  |  |  |  |  |  |
| **Racialized group<sup>c</sup>** |  |  |  |  |  |  |  |  |  |  |  |
| Latina/o | 1168 (56.0) | 1168 (100) | 0 (0.0) | 0 (0.0) | 0 (0.0) | 201 (26.4) | 322 (60.0) | 543 (84.3) | 775 (69.4) | 272 (40.7) | 121 (40.1) |
| White | 553 (26.5) | 0 (0.0) | 553 (100) | 0 (0.0) | 0 (0.0) | 395 (51.9) | 87 (16.2) | 50 (7.8) | 205 (18.4) | 273 (40.9) | 75 (24.8) |
| Black | 187 (9.0) | 0 (0.0) | 0 (0.0) | 187 (100) | 0 (0.0) | 100 (13.1) | 51 (9.5) | 25 (3.9) | 68 (6.1) | 84 (12.6) | 35 (11.6) |
| "Asian or other" | 178 (8.5) | 0 (0.0) | 0 (0.0) | 0 (0.0) | 178 (100) | 65 (8.5) | 77 (14.3) | 26 (4.0) | 68 (6.1) | 39 (5.8) | 71 (23.5) |
| **Gender** |  |  |  |  |  |  |  |  |  |  |  |
| Women | 2016 (97.3) | 1145 (98.5) | 524 (96.0) | 177 (94.7) | 170 (95.5) | 715 (95.0) | 527 (98.1) | 634 (98.8) | 1082 (97.5) | 651 (98.5) | 283 (93.7) |
| Men | 57 (2.7) | 17 (1.5) | 22 (4.0) | 10 (5.3) | 8 (4.5) | 38 (5.0) | 10 (1.9) | 8 (1.2) | 28 (2.5) | 10 (1.5) | 19 (6.3) |
| **Age (years)** |  |  |  |  |  |  |  |  |  |  |  |
| Mean (SD) | 42.6 (12.6) | 43.8 (11.5) | 37.6 (12.8) | 45.8 (13.3) | 47.7 (12.9) | 40.8 (14.3) | 45.7 (11.6) | 42.4 (10.6) | 44.2 (11.7) | 37.8 (12.7) | 47.7 (12.1) |
| Median (Min, Max) | 42 (19, 85) | 44 (19, 77) | 34 (19, 78) | 46 (19, 85) | 49 (20, 74) | 39 (19, 85) | 46 (20, 76) | 42 (19, 77) | 44 (19, 85) | 35 (19, 75) | 49 (20, 76) |
| [missing: n (%)]<sup>d</sup> | 33 (1.6) | 24 (2.1) | 6 (1.1) | 1 (0.5) | 2 (1.1) | 7 (0.9) | 9 (1.7) | 9 (1.4) | 15 (1.3) | 13 (1.9) | 5 (1.7) |
| **Formal educational attainment** |  |  |  |  |  |  |  |  |  |  |  |
| Less than 12 years | 713 (34.8) | 607 (53.0) | 44 (8.0) | 35 (19.7) | 27 (15.3) | 126 (16.8) | 184 (34.5) | 337 (53.3) | 504 (45.9) | 133 (20.2) | 76 (25.9) |
| ≥HSD (or equivalent) and <Bachelor's | 1081 (52.8) | 476 (41.5) | 370 (67.4) | 136 (76.4) | 99 (56.2) | 485 (64.5) | 286 (53.7) | 255 (40.3) | 527 (48.0) | 383 (58.2) | 171 (58.4) |
| Bachelor's degree or higher | 255 (12.4) | 63 (5.5) | 135 (24.6) | 7 (3.9) | 50 (28.4) | 141 (18.8) | 63 (11.8) | 40 (6.3) | 67 (6.1) | 142 (21.6) | 46 (15.7) |
| [missing: n (%)]<sup>d</sup> | 37 (1.8) | 22 (1.9) | 4 (0.7) | 9 (4.8) | 2 (1.1) | 9 (1.2) | 4 (0.7) | 12 (1.9) | 18 (1.6) | 10 (1.5) | 9 (3.0) |
| **Citizenship and immigration status<sup>b</sup>** |  |  |  |  |  |  |  |  |  |  |  |
| US citizen | 761 (39.2) | 201 (18.9) | 395 (74.2) | 100 (56.8) | 65 (38.7) | 761 (100.0) | 0 (0.0) | 0 (0.0) | 295 (28.3) | 333 (54.0) | 133 (47.0) |
| Documented immigrant | 537 (27.7) | 322 (30.2) | 87 (16.4) | 51 (29.0) | 77 (45.8) | 0 (0.0) | 537 (100.0) | 0 (0.0) | 302 (29.0) | 148 (24.0) | 87 (30.7) |
| Undocumented immigrant | 644 (33.2) | 543 (50.9) | 50 (9.4) | 25 (14.2) | 26 (15.5) | 0 (0.0) | 0 (0.0) | 644 (100.0) | 445 (42.7) | 136 (22.0) | 63 (22.3) |
| [missing: n (%)]<sup>d</sup> | 144 (6.9) | 102 (8.7) | 21 (3.8) | 11 (5.9) | 10 (5.6) | 0 (0.0) | 0 (0.0) | 0 (0.0) | 74 (6.6) | 51 (7.6) | 19 (6.3) |
| **Household characteristics** |  |  |  |  |  |  |  |  |  |  |  |
| **Income earner status** |  |  |  |  |  |  |  |  |  |  |  |
| Joint earner | 1023 (49.3) | 650 (56.0) | 248 (44.9) | 53 (28.6) | 72 (40.7) | 316 (41.7) | 283 (53.0) | 354 (55.2) | 591 (53.2) | 306 (46.1) | 126 (42.0) |
| Main earner | 416 (20.1) | 167 (14.4) | 126 (22.8) | 75 (40.5) | 48 (27.1) | 190 (25.1) | 103 (19.3) | 97 (15.1) | 207 (18.6) | 125 (18.8) | 84 (28.0) |
| Sole earner | 635 (30.6) | 343 (29.6) | 178 (32.2) | 57 (30.8) | 57 (32.2) | 251 (33.2) | 148 (27.7) | 190 (29.6) | 312 (28.1) | 233 (35.1) | 90 (30.0) |
| **Household economic insecurity** |  |  |  |  |  |  |  |  |  |  |  |
| Paid rent/mortgage and essential bill late | 634 (31.1) | 423 (37.1) | 100 (18.4) | 73 (40.1) | 38 (21.6) | 170 (22.8) | 161 (30.4) | 253 (40.3) | 379 (34.6) | 171 (26.2) | 84 (28.7) |
| Paid rent/mortgage or essential bill late | 502 (24.6) | 276 (24.2) | 135 (24.9) | 39 (21.4) | 52 (29.5) | 190 (25.4) | 134 (25.3) | 148 (23.6) | 246 (22.5) | 181 (27.7) | 75 (25.6) |
| Neither | 904 (44.3) | 440 (38.6) | 308 (56.7) | 70 (38.5) | 86 (48.9) | 387 (51.8) | 235 (44.3) | 227 (36.1) | 469 (42.9) | 301 (46.1) | 134 (45.7) |
| [missing: n (%)]<sup>d</sup> | 46 (2.2) | 29 (2.5) | 10 (1.8) | 5 (2.7) | 2 (1.1) | 14 (1.8) | 7 (1.3) | 16 (2.5) | 22 (2.0) | 15 (2.2) | 9 (3.0) |
| **N of people relying on financial support** |  |  |  |  |  |  |  |  |  |  |  |
| 0-1 | 871 (43.2) | 350 (30.6) | 366 (69.1) | 87 (50.6) | 68 (40.2) | 491 (66.6) | 192 (37.4) | 150 (23.8) | 366 (34.0) | 376 (57.9) | 129 (44.9) |
| 2 | 452 (22.4) | 292 (25.5) | 87 (16.4) | 41 (23.8) | 32 (18.9) | 117 (15.9) | 115 (22.4) | 185 (29.4) | 262 (24.3) | 123 (19.0) | 67 (23.3) |
| ≥3 | 691 (34.3) | 501 (43.8) | 77 (14.5) | 44 (25.6) | 69 (40.8) | 129 (17.5) | 206 (40.2) | 295 (46.8) | 450 (41.7) | 150 (23.1) | 91 (31.7) |
| [missing: n (%)]<sup>d</sup> | 72 (3.5) | 25 (2.1) | 23 (4.2) | 15 (8.0) | 9 (5.1) | 24 (3.2) | 24 (4.5) | 14 (2.2) | 38 (3.4) | 19 (2.8) | 15 (5.0) |
| **Employment and occupational characteristics** |  |  |  |  |  |  |  |  |  |  |  |
| **Years worked as DWer in the US** |  |  |  |  |  |  |  |  |  |  |  |
| Mean (SD) | 8.6 (7.4) | 8.7 (7.3) | 8.1 (7.4) | 10.0 (8.9) | 8.2 (6.7) | 9.3 (9.2) | 9.1 (7.1) | 7.6 (5.4) | 9.4 (8.0) | 7.5 (6.8) | 8.1 (6.6) |
| Median (Min, Max) | 6 (0.1, 60) | 6 (0.1, 50) | 5 (0.1, 38) | 9 (0.5, 60) | 7 (0.1, 32) | 6 (0.1, 60) | 8 (0.1, 37) | 6 (0.1, 31) | 7 (0.1, 60) | 5 (0.1, 45) | 6 (0.2, 38) |
| [missing: n (%)]<sup>d</sup> | 46 (2.2) | 38 (3.3) | 5 (0.9) | 1 (0.5) | 2 (1.1) | 10 (1.3) | 9 (1.7) | 22 (3.4) | 34 (3.0) | 8 (1.2) | 4 (1.3) |
| **Main DW occupation** |  |  |  |  |  |  |  |  |  |  |  |
| Housecleaning | 1116 (53.5) | 775 (66.4) | 205 (37.1) | 68 (36.4) | 68 (38.2) | 295 (38.8) | 302 (56.2) | 445 (69.1) | 1116 (100) | 0 (0.0) | 0 (0.0) |
| Child care | 668 (32.0) | 272 (23.3) | 273 (49.4) | 84 (44.9) | 39 (21.9) | 333 (43.8) | 148 (27.6) | 136 (21.1) | 0 (0.0) | 668 (100) | 0 (0.0) |
| Adult care | 302 (14.5) | 121 (10.4) | 75 (13.6) | 35 (18.7) | 71 (39.9) | 133 (17.5) | 87 (16.2) | 63 (9.8) | 0 (0.0) | 0 (0.0) | 302 (100) |
| **Live-in DWer** | 245 (11.7) | 142 (12.2) | 35 (6.3) | 15 (8.0) | 53 (29.8) | 51 (6.7) | 93 (17.3) | 83 (12.9) | 102 (9.1) | 80 (12.0) | 63 (20.9) |
| **Contract, any DW employer, last 12 mo.** |  |  |  |  |  |  |  |  |  |  |  |
| No | 1098 (53.0) | 670 (57.8) | 225 (40.8) | 114 (61.0) | 89 (51.4) | 351 (46.6) | 304 (56.9) | 369 (57.7) | 636 (57.4) | 318 (48.0) | 144 (48.2) |
| Yes | 948 (45.8) | 469 (40.5) | 325 (59.0) | 73 (39.0) | 81 (46.8) | 398 (52.8) | 222 (41.6) | 261 (40.8) | 456 (41.2) | 341 (51.4) | 151 (50.5) |
| Don't know | 24 (1.2) | 20 (1.7) | 1 (0.2) | 0 (0.0) | 3 (1.7) | 5 (0.7) | 8 (1.5) | 9 (1.4) | 16 (1.4) | 4 (0.6) | 4 (1.3) |
| **Working hours, main DW employer, last week** |  |  |  |  |  |  |  |  |  |  |  |
| Mean (SD) | 28.5 (21.4) | 26.2 (20.2) | 27.3 (19.3) | 32.7 (21.0) | 43.7 (28.9) | 25.9 (18.8) | 33.2 (23.6) | 27.2 (21.6) | 22.5 (19.7) | 34.2 (18.1) | 38.3 (26.9) |
| Median (Min, Max) | 22.5 (1.5, 168.0) | 20.3 (2, 120.0) | 23.0 (1.5, 113.2) | 30.0 (4.0, 119.0) | 42.0 (3.5, 168.0) | 20.5 (1.5, 168.0) | 28.0 (3.0, 160.5) | 20.2 (3.8, 118.5) | 15.2 (2.0, 119.0) | 33.2 (1.5, 118.5) | 31.0 (4.0, 168.0) |
| [missing: n (%)]<sup>d</sup> | 30 (1.4) | 6 (0.5) | 13 (2.4) | 2 (1.1) | 9 (5.1) | 5 (0.7) | 9 (1.7) | 13 (2.0) | 14 (1.3) | 6 (0.9) | 10 (3.3) |
| **Number of DW employers, last mo.** |  |  |  |  |  |  |  |  |  |  |  |
| Mean (SD) | 2.0 (2.7) | 2.1 (2.7) | 2.2 (3.2) | 1.3 (0.8) | 1.5 (1.4) | 2.0 (2.8) | 1.8 (2.5) | 2.2 (2.8) | 2.5 (3.4) | 1.4 (1.2) | 1.2 (0.8) |
| Median (Min, Max) | 1.0 (0, 35) | 1.0 (0, 30) | 1.0 (0, 35) | 1.0 (0, 5) | 1.0 (0, 10) | 1.0 (0, 35) | 1.0 (0, 35) | 1.0 (0, 30) | 1.0 (0, 35) | 1.0 (0, 11) | 1.0 (0, 8) |
| [missing: n (%)]<sup>d</sup> | 59 (2.8) | 26 (2.2) | 19 (3.4) | 9 (4.8) | 5 (2.8) | 26 (3.4) | 15 (2.8) | 12 (1.9) | 23 (2.1) | 21 (3.1) | 15 (5.0) |
| **Workplace hazards, last 12 months, any domestic work employer** |  |  |  |  |  |  |  |  |  |  |  |
| **Paid less than agreed** | 242 (11.6) | 119 (10.2) | 78 (14.1) | 13 (7.0) | 32 (18.1) | 89 (11.7) | 65 (12.1) | 77 (12.0) | 133 (11.9) | 77 (11.5) | 32 (10.7) |
| **Paid late** | 489 (23.5) | 264 (22.7) | 155 (28.0) | 27 (14.5) | 43 (24.2) | 169 (22.2) | 125 (23.3) | 163 (25.4) | 262 (23.5) | 160 (24.0) | 67 (22.2) |
| **Paid with bad check** | 70 (3.4) | 39 (3.4) | 15 (2.7) | 7 (3.8) | 9 (5.1) | 26 (3.4) | 20 (3.7) | 18 (2.8) | 37 (3.3) | 17 (2.6) | 16 (5.3) |
| **Charged for something broken or lost** | 120 (5.8) | 70 (6.0) | 32 (5.8) | 8 (4.3) | 10 (5.6) | 31 (4.1) | 30 (5.6) | 52 (8.1) | 92 (8.3) | 18 (2.7) | 10 (3.3) |
| **Deductions from pay without consent** | 110 (5.3) | 58 (5.0) | 35 (6.3) | 8 (4.3) | 9 (5.1) | 35 (4.6) | 31 (5.8) | 38 (6.0) | 63 (5.7) | 34 (5.1) | 13 (4.3) |
| **Pressured to work more than scheduled hours** | 391 (18.8) | 187 (16.1) | 136 (24.6) | 28 (15.1) | 40 (22.5) | 145 (19.1) | 107 (20.0) | 122 (19.0) | 207 (18.6) | 128 (19.2) | 56 (18.5) |
| **Verbally abused or yelled at** | 328 (15.8) | 192 (16.5) | 83 (15.0) | 19 (10.2) | 34 (19.1) | 98 (12.9) | 96 (18.0) | 120 (18.6) | 172 (15.5) | 82 (12.3) | 74 (24.5) |
| **Threatened** | 122 (5.9) | 70 (6.0) | 26 (4.7) | 6 (3.2) | 20 (11.3) | 26 (3.4) | 46 (8.6) | 45 (7.0) | 68 (6.2) | 30 (4.5) | 24 (7.9) |
| **Used immigration status to threaten** | 71 (3.4) | 51 (4.4) | 6 (1.1) | 2 (1.1) | 12 (6.8) | 1 (0.1) | 24 (4.5) | 44 (6.8) | 43 (3.9) | 20 (3.0) | 8 (2.6) |
| **Kept immigration papers** | 12 (0.6) | 5 (0.4) | 1 (0.2) | 2 (1.1) | 4 (2.3) | 0 (0.0) | 8 (1.5) | 4 (0.6) | 8 (0.7) | 4 (0.6) | 0 (0.0) |
| **Called insulting names or racial slurs** | 159 (7.7) | 98 (8.5) | 26 (4.7) | 14 (7.5) | 21 (11.9) | 41 (5.4) | 44 (8.3) | 69 (10.7) | 89 (8.0) | 33 (5.0) | 37 (12.3) |
| **Pushed or shoved** | 49 (2.4) | 20 (1.7) | 20 (3.6) | 4 (2.2) | 5 (2.8) | 17 (2.2) | 13 (2.4) | 15 (2.3) | 19 (1.7) | 13 (2.0) | 17 (5.6) |
| **Physically hurt or attacked** | 37 (1.8) | 18 (1.6) | 15 (2.7) | 1 (0.5) | 3 (1.7) | 14 (1.8) | 11 (2.1) | 11 (1.7) | 11 (1.0) | 11 (1.7) | 15 (5.0) |
| [missing: n (%)]<sup>d</sup> | 25 (1.2) | 16 (1.4) | 4 (0.7) | 3 (1.6) | 2 (1.1) | 8 (1.1) | 7 (1.3) | 6 (0.9) | 18 (1.6) | 5 (0.7) | 2 (0.7) |
| **Did heavy lifting or other strenuous activities** | 833 (40.0) | 459 (39.3) | 251 (45.8) | 47 (25.1) | 76 (42.7) | 293 (38.7) | 196 (36.5) | 292 (45.3) | 516 (46.4) | 185 (27.8) | 132 (43.9) |
| **Climbed to clean** | 871 (41.9) | 583 (50.1) | 192 (35.0) | 39 (20.9) | 57 (32.0) | 241 (31.8) | 224 (41.8) | 340 (53.0) | 672 (60.4) | 130 (19.5) | 69 (23.0) |
| **Cared for someone with a contagious illness** | 223 (10.8) | 48 (4.1) | 143 (26.1) | 19 (10.2) | 13 (7.3) | 141 (18.7) | 46 (8.6) | 31 (4.8) | 41 (3.7) | 146 (22.0) | 36 (12.0) |
| **Worked long hours with no breaks** | 745 (35.9) | 375 (32.2) | 257 (46.9) | 48 (25.7) | 65 (36.5) | 271 (35.8) | 203 (37.8) | 230 (35.9) | 413 (37.2) | 245 (36.8) | 87 (28.9) |
| **Required to work on knees** | 736 (35.5) | 434 (37.3) | 223 (40.7) | 29 (15.6) | 50 (28.2) | 231 (30.5) | 183 (34.1) | 268 (41.9) | 538 (48.6) | 141 (21.2) | 57 (18.9) |
| **Worked with toxic cleaning supplies** |  |  |  |  |  |  |  |  |  |  |  |
| Yes | 963 (46.7) | 663 (57.2) | 191 (35.4) | 55 (30.1) | 54 (30.5) | 244 (32.8) | 271 (50.8) | 375 (58.5) | 733 (66.5) | 140 (21.2) | 90 (30.1) |
| No | 1057 (51.3) | 482 (41.6) | 328 (60.7) | 126 (68.9) | 121 (68.4) | 483 (65.0) | 252 (47.3) | 254 (39.6) | 336 (30.5) | 514 (78.0) | 207 (69.2) |
| Don't know<sup>e</sup> | 40 (1.9) | 15 (1.3) | 21 (3.9) | 2 (1.1) | 2 (1.1) | 16 (2.2) | 10 (1.9) | 12 (1.9) | 33 (3.0) | 5 (0.8) | 2 (0.7) |
| [missing: n (%)]<sup>d</sup> | 26 (1.2) | 8 (0.7) | 13 (2.4) | 4 (2.1) | 1 (0.6) | 18 (2.4) | 4 (0.7) | 3 (0.5) | 14 (1.3) | 9 (1.3) | 3 (1.0) |
| **Health status** |  |  |  |  |  |  |  |  |  |  |  |
| **Work-related back injury, last 12 mo.** | 558 (26.8) | 366 (31.4) | 116 (21.1) | 39 (20.9) | 37 (20.9) | 160 (21.1) | 157 (29.3) | 213 (33.1) | 345 (31.0) | 124 (18.6) | 89 (29.5) |
| **Work-related illness, last 12 mo.** | 680 (32.6) | 394 (33.8) | 212 (38.5) | 37 (19.8) | 37 (20.8) | 232 (30.6) | 152 (28.3) | 240 (37.3) | 333 (29.9) | 268 (40.2) | 79 (26.2) |
| **Fair-to-poor self-rated health, last 12 mo.** | 759 (37.1) | 559 (48.9) | 111 (20.4) | 31 (16.8) | 58 (33.3) | 185 (24.6) | 210 (39.8) | 304 (48.3) | 477 (43.6) | 163 (24.8) | 119 (40.5) |
| [missing: n (%)]<sup>d</sup> | 40 (1.9) | 24 (2.1) | 10 (1.8) | 2 (1.1) | 4 (2.2) | 9 (1.2) | 10 (1.9) | 14 (2.2) | 22 (2) | 10 (1.5) | 8 (2.6) |
| **No health insurance** | 1258 (60.8) | 855 (74.0) | 213 (38.7) | 102 (54.5) | 88 (49.7) | 264 (34.9) | 343 (64.0) | 538 (84.3) | 774 (70.0) | 330 (49.7) | 154 (51.3) |

Abbreviations: Doc = documented; DW = domestic work; DWer = domestic worker; HSD = high school diploma; mo = month(s); N = number; SD = standard deviation; Undoc = undocumented; US = United States
<sup>a</sup> Los Angeles, CA; San Francisco, CA; San Jose, CA; San Diego, CA; Denver, CO; Washington DC; Atlanta, GA; Miami, FL; Chicago, IL; Boston, MA; New York, NY; Houston, TX; San Antonio, TX; Seattle, WA
<sup>b</sup> Six participants were originally coded as having "Other" immigration status. Those described—in an accompanying text field—as "non-resident" (n = 1); "immigrant" (n = 1); "RI" (n = 1); and no description (n = 2) were coded as "missing;" the one described as "work visa" (n = 1) was coded as visa-holder (documented immigrant).
<sup>c</sup> The categories shown are the categories employed in the dataset.
<sup>d</sup> Percent missing based on total; otherwise, distributions are based on observed cases only. Missingness not shown for variables with <1% (n < 21) missing data in the total sample.
<sup>e</sup> For variables that include a response option for "Don't know," rows for "Yes," "No," and "Don't know" are shown.

Investigators employed participatory methods whereby organizers from 34 NDWA-affiliated community organizations and 190 DWers collaborated on survey design, fielding, and analysis. Because of the secluded, dispersed nature of DW, surveyors recruited participants at public places (i.e., parks, transportation hubs, churches, shopping centers) and used chain-referral of potential participants. Surveys were conducted in nine languages (Cantonese, English, Haitian Creole, Mandarin, Nepali, Polish, Portuguese, Spanish, Tagalog). DWers were eligible to participate if they worked in a private home during the previous week for ≥6 hours as a housecleaning, childcare, or adult care worker; were paid directly by an employing family member; were ≥18 years old; lived in a selected city; and were not members of organizations that advocated for workers’ right—so as to avoid bias potentially related to “participants who had more knowledge about exercising their employment rights.”[18] The Harvard T.H. Chan School of Public Health Institutional Review Board determined our secondary analyses of the NDWA-UIC CUED data was Not Human Subjects Research (IRB21-0855).

### Workplace hazards

In the NDWA-UIC CUED survey, participants self-reported exposure (yes/no) to six economic (e.g., paid late), nine social (e.g., verbally abused), and six occupational (e.g., heavy lifting) hazards (n=21 variables) at any DW job in the previous 12 months (**Table 1**). The option to respond “don’t know” was available only for the “worked with toxic cleaning supplies” variable. We coded as missing the few participants who responded using this option (n=40 [1.9%]). Although the NDWA-UIC CUED survey asked about exposure to workplace sexual harassment and sexual assault, we excluded these variables from the analysis due to severe underreporting,[20,21] thereby reducing the number of exposure variables to 19 (for more details, see **Supplemental Methods**).

### Health outcomes

We used self-reported data on whether DWers reported experiencing back injury (“Back injuries, including pulled back muscles”) and, separately, illness (“Contracted an illness, such as the flu”) while working at any job as a DWer in the previous 12 months. The survey also inquired about participants’ self-rated health in the previous 12 months (1=Excellent, 2=Very good, 3=Good, 4=Fair, 5=Poor), which we categorized as fair-to-poor self-rated health (FPSRH; 1=Fair-to-poor, 0=Excellent, very good, or good). These three outcomes were commonly experienced by DWers in the sample (back injury: 26.8%; illness: 32.6%; FPSRH: 37.1%).

### Covariates

The following covariates were included in all health outcome models: racialized group (collected as: “White,” “Latina/o,” “Black,” or “Asian or other,” which participants could, but in no cases did, multiply select), citizenship and immigration status, gender, age, education, main DW occupation, live-in status, years worked as a DWer in the US, hours worked last week for one’s main DW employer, number of DW employers, contract status, number of people relying on the participant’s financial support, income earner status, and household economic insecurity. Covariate selection was informed by our use of ecosocial theory,[3, 16, 17] prior research identifying predictors of workplace exposures and/or health among DWers, observed variation in DWer exposures by covariates, and empirical considerations regarding collinearity and model complexity (for more details, see **Supplemental Methods**).

### Statistical analyses

We used R (Version 4.1.3, R Foundation for Statistical Computing, Vienna, Austria) to conduct all statistical analyses except latent class analysis, which was conducted in LatentGOLD (Version 6.0, Statistical Innovations, Inc., Arlington, VA).[22, 23] All tests for statistical significance were two-sided. We first descriptively quantified the distribution of—and examined relationships between—variables in the dataset, including regarding missingness (e.g., **Figure S1**).

We then defined exposure to workplace hazards using the four approaches described below and fit models to estimate associations between exposure and each health outcome (**Table 2**). All health outcome models adjusted for the aforementioned covariates, and used city fixed effects to account for all unobserved city-specific, time-invariant characteristics that may confound exposure-outcome associations.[24] Hausman tests (i.e., tests of the adequacy of city random effects models) indicated the need for city fixed effects to address endogeneity in models for each of the three health outcomes.[25]

**Table 2.**
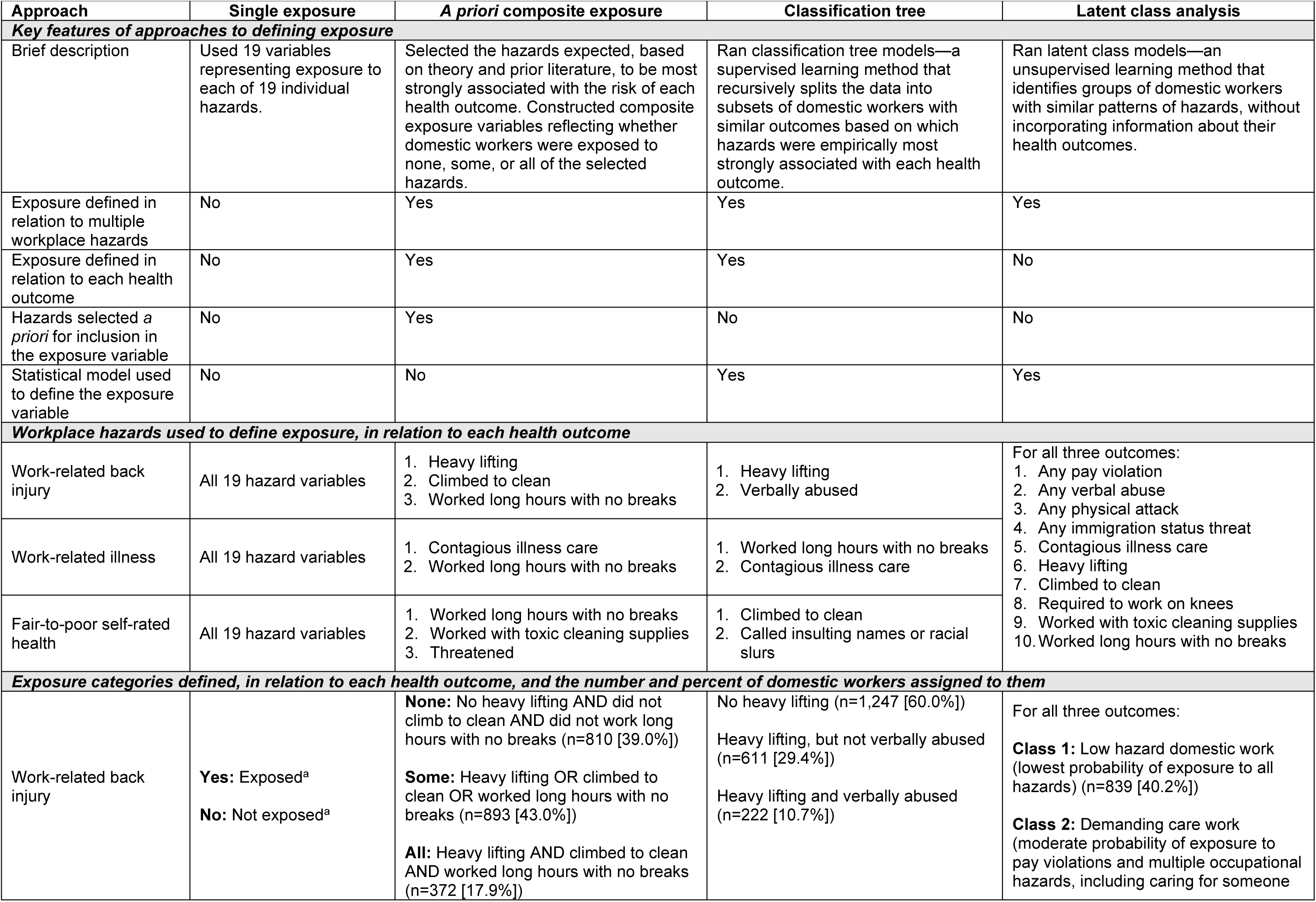
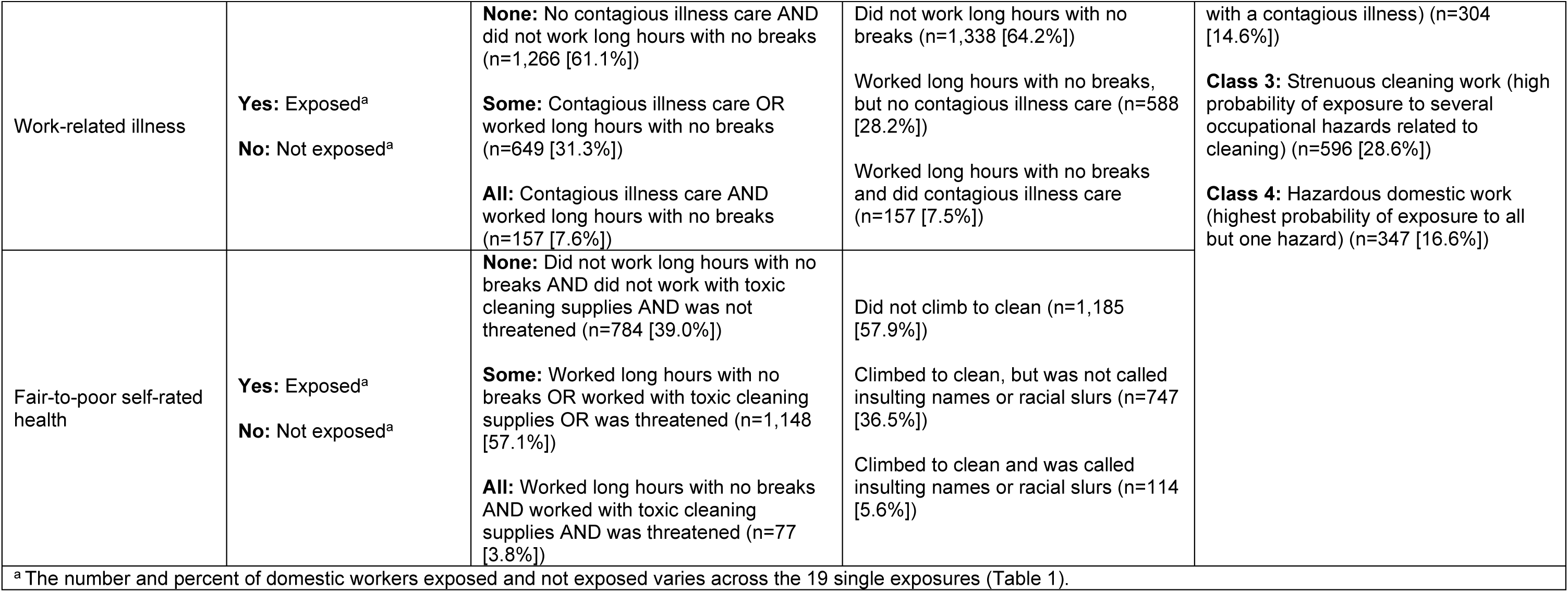
**Overview of approaches to defining exposure: National Domestic Workers Alliance and University of Illinois Chicago Center for Urban Economic Development data, 14 cities, United States, 2011-2012 (N = 2**,**086)**.

|  |  |  |  |  |
| --- | --- | --- | --- | --- |
| Approach | Single exposure | A priori composite exposure | Classification tree | Latent class analysis |
| Key features of approaches to defining exposure |  |  |  |  |
| Brief description | Used 19 variables representing exposure to each of 19 individual hazards. | Selected the hazards expected, based on theory and prior literature, to be most strongly associated with the risk of each health outcome. Constructed composite exposure variables reflecting whether domestic workers were exposed to none, some, or all of the selected hazards. | Ran classification tree models—a supervised learning method that recursively splits the data into subsets of domestic workers with similar outcomes based on which hazards were empirically most strongly associated with each health outcome. | Ran latent class models—an unsupervised learning method that identifies groups of domestic workers with similar patterns of hazards, without incorporating information about their health outcomes. |
| Exposure defined in relation to multiple workplace hazards | No | Yes | Yes | Yes |
| Exposure defined in relation to each health outcome | No | Yes | Yes | No |
| Hazards selected a priori for inclusion in the exposure variable | No | Yes | No | No |
| Statistical model used to define the exposure variable | No | No | Yes | Yes |
| Workplace hazards used to define exposure, in relation to each health outcome |  |  |  |  |
| Work-related back injury | All 19 hazard variables | 1. Heavy lifting<br>2. Climbed to clean<br>3. Worked long hours with no breaks | 1. Heavy lifting<br>2. Verbally abused | For all three outcomes:<br>1. Any pay violation<br>2. Any verbal abuse<br>3. Any physical attack<br>4. Any immigration status threat<br>5. Contagious illness care<br>6. Heavy lifting<br>7. Climbed to clean<br>8. Required to work on knees<br>9. Worked with toxic cleaning supplies<br>10. Worked long hours with no breaks |
| Work-related illness | All 19 hazard variables | 1. Contagious illness care<br>2. Worked long hours with no breaks | 1. Worked long hours with no breaks<br>2. Contagious illness care |  |
| Fair-to-poor self-rated health | All 19 hazard variables | 1. Worked long hours with no breaks<br>2. Worked with toxic cleaning supplies<br>3. Threatened | 1. Climbed to clean<br>2. Called insulting names or racial slurs |  |
| Exposure categories defined, in relation to each health outcome, and the number and percent of domestic workers assigned to them |  |  |  |  |
| Work-related back injury | Yes: Exposed<sup>a</sup><br>No: Not exposed<sup>a</sup> | None: No heavy lifting AND did not climb to clean AND did not work long hours with no breaks (n=810 [39.0%])<br>Some: Heavy lifting OR climbed to clean OR worked long hours with no breaks (n=893 [43.0%])<br>All: Heavy lifting AND climbed to clean AND worked long hours with no breaks (n=372 [17.9%]) | No heavy lifting (n=1,247 [60.0%])<br>Heavy lifting, but not verbally abused (n=611 [29.4%])<br>Heavy lifting and verbally abused (n=222 [10.7%]) | For all three outcomes:<br>Class 1: Low hazard domestic work (lowest probability of exposure to all hazards) (n=839 [40.2%])<br>Class 2: Demanding care work (moderate probability of exposure to pay violations and multiple occupational hazards, including caring for someone |
| Work-related illness | **Yes:** Exposed<sup>a</sup><br>**No:** Not exposed<sup>a</sup> | **None:** No contagious illness care AND did not work long hours with no breaks (n=1,266 [61.1%])<br>**Some:** Contagious illness care OR worked long hours with no breaks (n=649 [31.3%])<br>**All:** Contagious illness care AND worked long hours with no breaks (n=157 [7.6%]) | Did not work long hours with no breaks (n=1,338 [64.2%])<br>Worked long hours with no breaks, but no contagious illness care (n=588 [28.2%])<br>Worked long hours with no breaks and did contagious illness care (n=157 [7.5%]) | with a contagious illness) (n=304 [14.6%])<br>**Class 3:** Strenuous cleaning work (high probability of exposure to several occupational hazards related to cleaning) (n=596 [28.6%])<br>**Class 4:** Hazardous domestic work (highest probability of exposure to all but one hazard) (n=347 [16.6%]) |
| Fair-to-poor self-rated health | **Yes:** Exposed<sup>a</sup><br>**No:** Not exposed<sup>a</sup> | **None:** Did not work long hours with no breaks AND did not work with toxic cleaning supplies AND was not threatened (n=784 [39.0%])<br>**Some:** Worked long hours with no breaks OR worked with toxic cleaning supplies OR was threatened (n=1,148 [57.1%])<br>**All:** Worked long hours with no breaks AND worked with toxic cleaning supplies AND was threatened (n=77 [3.8%]) | Did not climb to clean (n=1,185 [57.9%])<br>Climbed to clean, but was not called insulting names or racial slurs (n=747 [36.5%])<br>Climbed to clean and was called insulting names or racial slurs (n=114 [5.6%]) |  |

<sup>a</sup> The number and percent of domestic workers exposed and not exposed varies across the 19 single exposures (Table 1).

Because all three health outcomes were non-rare in the sample, models were fit using log Poisson regression, with parameter estimates interpretable as log risk ratios (RR).[26] All health outcome models employed multiply imputed covariates and health outcome data. We used multiple imputation using chained equations to create 10 imputed datasets under the Missing-At-Random (MAR) assumption.[27] We included in the imputation model all variables to be included in the health outcome models and additional variables that may have otherwise explained nonresponse or variance in the data (for more details, see **Supplemental Methods**). All 95% confidence intervals are based on cluster robust standard errors, clustered at the city level.

#### Single exposure

We first estimated the RR of each health outcome associated with exposure to the 19 individual hazards by including them singly in separate models. Because this involved testing multiple dependent hypotheses, we used Benjamini and Yekutieli’s (2001) approach to control the False Discovery Rate.[28]

#### *A priori* composite exposure

To examine how exposure to multiple hazards was associated with health—without relying on complex exposure modeling strategies—we selected the hazards we expected would be most strongly associated with each health outcome. For work-related back injury, work-related illness, and FPSRH, respectively, these were: 1) heavy lifting, climbing to clean, and working long hours with no breaks, 2) caring for someone with a contagious illness, and working long hours with no breaks, and 3) working long hours with no breaks, working with toxic cleaning supplies, and being threatened (for more details, see **Supplemental Methods**). We constructed composite exposure variables reflecting whether DWers were exposed to none, some, or all of these selected hazards. We estimated the RR of each health outcome associated with experiencing some (versus none) or all (versus none) of the selected hazards for that outcome.

#### Classification trees

We employed classification trees to empirically identify the combinations of workplace exposures most strongly associated with each outcome. Classification trees are a supervised learning method that identifies groups of individuals with similar outcomes based on their patterns of responses to multiple observed variables.[14, 29] This method identifies the single hazard that best predicts the outcome, splits the data into two groups according to exposure to that hazard, and recursively repeats this process with each identified subgroup until splitting by additional variables no longer improves the results (for more details, see **Supplemental Methods**). Thus, each resulting classification tree was composed of “branches” representing exposures, on the basis of which DWers were grouped into “leaves,” representing the final groupings of DWers. Using these final groupings, we estimated the RR of each outcome associated with experiencing each exposure pattern (versus the least exposed group) for that outcome.

#### Latent class analysis

We used latent class analysis (LCA) to identify groups of DWers with distinct patterns of exposure to workplace hazards, regardless of the health outcomes they reported. LCA is an unsupervised learning method (i.e., no outcome variables) that identifies groups of individuals, called latent classes, based on the clustering of their responses to multiple observed variables.[30, 31] This method uses the statistical associations between observed variables to learn about the structure of an assumed underlying categorical latent variable with an unknown number of classes. We determined the number of latent classes by comparing the fit, classification certainty, identifiability, and interpretability of multiple fitted LCA models with differing number of latent classes.[12] We initially included all 19 workplace hazard variables in these models. However, violations of the LCA conditional independence assumption—which states that membership in the latent classes explains all associations between variables—can arise due to redundancies between observed variables and lead to the selection of too many latent classes.[32] Thus, we monitored for violations of this assumption, considered combining or dropping redundant hazard variables as needed, and reran the LCA models with this reduced set of variables before deciding how many latent classes to retain. We estimated the RR of each health outcome associated with membership in each latent class (versus the least exposed class). See Wright et al [12] and the **Supplemental Methods** for more details on our latent class approach.

## RESULTS

Descriptive characteristics of the 2,086 informally employed DWers surveyed are presented in **Table 1**. Regarding single workplace hazards, more than 15 percent of DWers reported being verbally abused or yelled at (15.8%), pressured to work more than their scheduled hours (18.8%), paid late (23.5%), required to work on their knees (35.5%), working long hours with no breaks (35.9%), doing heavy lifting (40.0%), climbing to clean (41.9%), and working with toxic cleaning supplies (46.7%). Exposure to single hazards varied by racialized group, citizenship and immigration status, and main DW occupation. The percent of DWers who reported a work-related back injury, work-related illness, and FPSRH, respectively, equaled 26.8%, 32.6%, and 37.1%.

Results from the classification trees are presented in **Figure 1, Figure S2**, and **Figure S3**. Classification trees identified “heavy lifting” and “verbally abused” as the variables most strongly associated with work-related back injury, yielding exposure categories for “No heavy lifting” (n=1,247 [60.0%]), “Heavy lifting, but no verbal abuse” (n=611 [29.4%]), and “Heavy lifting and verbal abuse” (n=222 [10.7%]). For work-related illness, classification trees yielded exposure categories for “Did not work long hours with no breaks” (n=1,338 [64.2%]), “Worked long hours, but did no contagious illness care” (n=588 [28.2%]), and “Worked long hours and did contagious illness care (n=157 [7.5%]). For FPSRH, classification trees identified “Did not climb to clean” (n=1,185 [57.9%]), “Climbed to clean, but was not called insulting names or racial slurs” (n=747 [36.5%]), and “Climbed to clean and was called insulting names or racial slurs” (n=114 [5.6%]). Classification trees yielded similar exposure categories to *a priori* composite variables (**Table 2**). For example, *a priori* selection and classification trees both identified “contagious illness care” and “worked long hours with no breaks” as the most important variables for predicting work-related illness.

**Figure 1.**
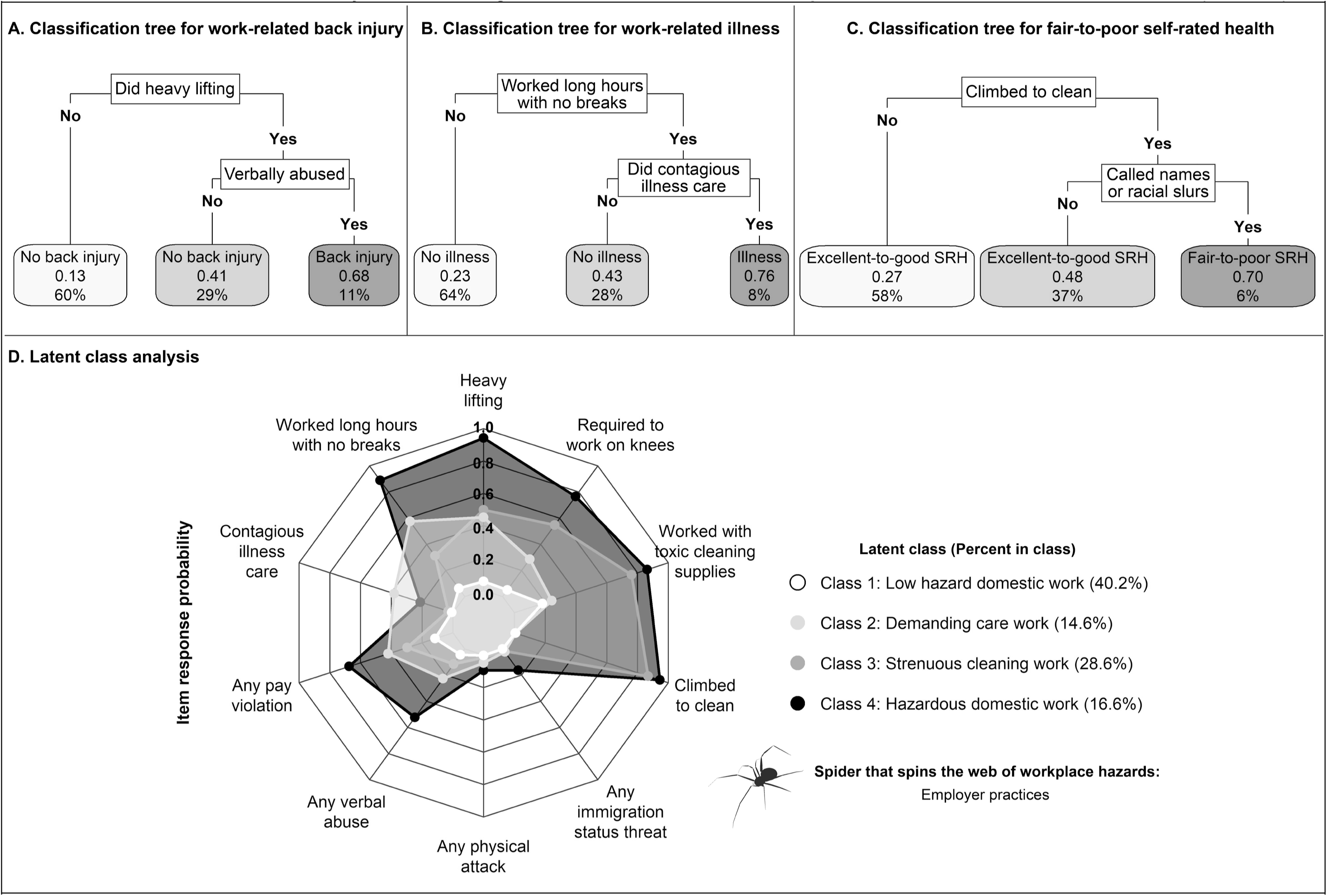
**Results from classification tree and latent class approaches to defining domestic workers’ exposures to multiple workplace hazards: National Domestic Workers Alliance and University of Illinois Chicago Center for Urban Economic Development data, 14 cities, United States, 2011-2012 (N = 2**,**086)**. Regarding classification trees (Figure 1A-C), figures show the final, pruned classification trees. Unpruned classification trees are provided in Figure S2. Each plot shows, in order of the elements of the figure from top to bottom: the name of the splitting variable (black textboxes), the splitting variable categories that determine how individuals are sorted based on that splitting variable (i.e., “No” exposure to a given hazard; “Yes” exposed to that hazard), and the final groups of individuals based on their responses to all splitting variables (shaded red textboxes). Within each final shaded red textbox is: a) the predicted health outcome for members of that group (e.g., “No illness,” “Illness”), b) the predicted probability of experiencing the health outcome of interest (e.g., the predicted probability of experiencing back injury among members of the first group in the work-related back injury classification tree equals 0.13), and c) the percent of the total sample (n=2,086) assigned to that leaf of the tree (e.g., 60% of the total sample is assigned to the first group in the work-related back injury classification tree). Regarding the latent class analysis, Figure 1D plots the estimated item response probability (range: 0-1) among domestic workers assigned to each latent class (i.e., the probability of being exposed to each hazard, conditional on being a member of a given class). Numerical values of the item response probabilities and their robust standard errors are provided in Table S1. Included below the legend for this “spider plot,” which depicts the web of workplace hazards domestic workers jointly experienced, is the metaphorical spider that spins this web: employer practices.[1-3, 16-17]

Latent class analysis suggested that four latent classes—for which we developed concise summary labels—best represented the patterns of workplace hazard exposure in the data (**Figure 1, Table S1**). Members of Class 1, which we labeled as “Low hazard domestic work,” had the lowest probability of exposure to all 10 hazards included in the final model (all item response probabilities [IRP], representing the probability of exposure among members of a given class, <0.2) (n=839 [40.2%]). Members of Class 2, labeled “Demanding care work” (n=304 [14.6%]), had moderate probability of exposure to pay violations (IRP=0.42), heavy lifting (IRP=0.45), working long hours with no breaks (IRP=0.57), and contagious illness care (IRP=0.38). Members of Class 3, labeled “Strenuous cleaning work” (n=596 [28.6%]), had high probability of exposure to several cleaning-related occupational hazards (e.g., IRP_climbed to clean_=0.87; IRP_toxic cleaning supplies_=0.76). Members of Class 4, labeled “Hazardous domestic work” (n=347 [16.6%]), had the highest probability of exposure to all but one hazard (contagious illness care).

Results of city fixed effects models indicated, first, that—across all four approaches to exposure definition—exposure to workplace hazards was associated with the risk of each health outcome, after adjusting for selected covariates (**Figure 2**). Across all three health outcomes, the single exposure approach produced the smallest (though often still notable and statistically significant, after adjusting for multiple testing [**Figure S4**]) estimates of the RR, whereas the *a priori* composite exposure, classification tree, and latent class analysis approaches performed comparably to one another. For example, for work-related back injury, the estimated RR associated with heavy lifting (the single exposure with the largest RR), exposure to all three hazards selected *a priori* (did heavy lifting, climbed to clean, worked long hours) versus none, exposure to the two hazards identified by classification trees (heavy lifting, verbally abused) versus “No heavy lifting,” and membership in the “hazardous domestic work” latent class versus the least-exposed “low hazard domestic work” class were, respectively, 3.4 (95% CI 2.7 to 4.1); 6.5 (95% CI 4.8 to 8.7); 4.4 (95% CI 3.6 to 5.3), and 6.6 (95% CI 4.6 to 9.4). Additional results from health outcome models are presented in **Table S2** (complete results from fully-adjusted models using the *a priori*, classification tree, and latent class approaches), **Table S3** (sensitivity analyses for unmeasured confounding), and **Figure S5** (stepwise inclusion of covariates in models). Descriptive figures further characterized the individual, household, and occupational characteristics of DWers in each exposure category, as defined according to each approach (**Figure S6**).

**Figure 2.**
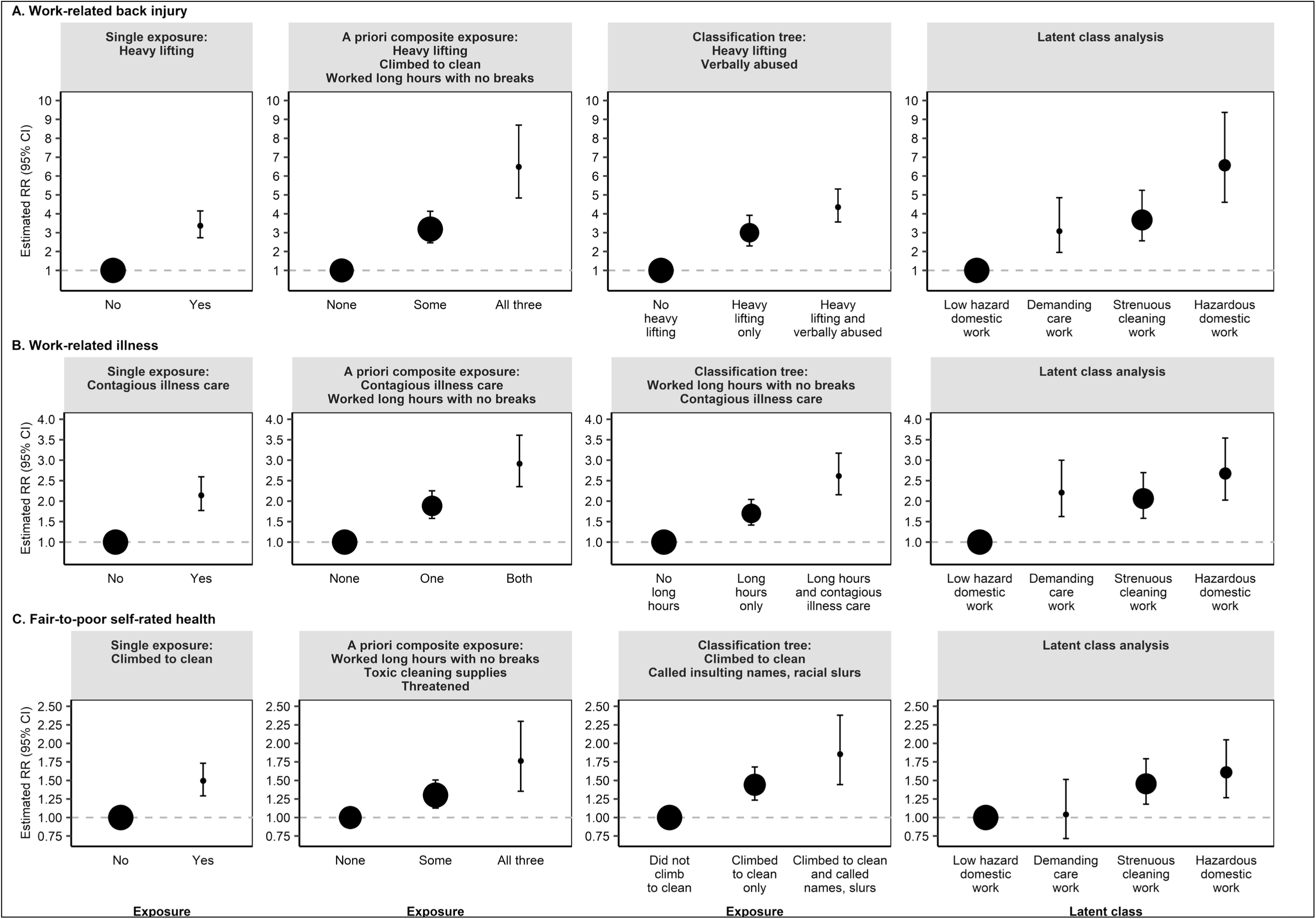
**Estimated risk ratios and 95% confidence intervals of work-related back injury, work-related illness, and fair-to-poor self-rated health—and relative size of exposure categories— from fully-adjusted city fixed effects models using four approaches to defining exposure: National Domestic Workers Alliance and University of Illinois Chicago Center for Urban Economic Development data, 14 cities, United States, 2011-2012 (N = 2**,**086)**. Abbreviations: CI = confidence interval; RR = risk ratio. The size of each point is equal to the square root of the number of domestic workers assigned to the designated exposure category for each approach. All models were adjusted for the following covariates: racialized group, citizenship and immigration status, gender, age, formal educational attainment, main domestic work occupation, live-in status, years worked as a domestic worker in the United States, hours worked last week for one’s main domestic work employer, number of domestic work employers, had a contract with any of their domestic work employers in the last 12 months, number of people relying on the participant’s financial support, income earner status, and household economic insecurity. All confidence intervals are based on cluster robust standard errors, clustered at the city level. For the single exposure approach, we plotted results for the one hazard (out of 19) with the largest and most reliable estimate of the risk ratio with each health outcome.

## DISCUSSION

Results from our novel, descriptive study indicate that—net of key individual, household, and occupational characteristics as well as city fixed effects—informally employed US DWers exposed to different combinations of workplace hazards experienced divergent health profiles. Compared to the three approaches used to characterize workers’ joint exposures, the single exposures approach underestimated the risk associated with workers’ lived experiences of multiple hazards. Compared to the *a priori* composite exposures, the classification tree approach estimated slightly smaller RR, reflecting larger exposure referent groups with greater prevalence of the outcome. Our findings also suggest that an *a priori* composite exposure approach—when informed explicitly by theory and subject matter expertise—can provide an accessible tool for defining exposure categories that meaningfully capture risk of a given outcome among DWers. Finally, despite being the only approach that did not incorporate information about DWers’ health outcomes in constructing exposure categories, the latent class approach produced RR estimates, across all three outcomes, of similar magnitude to those observed using the *a priori* and classification tree approaches.

The strengths and limitations of our study must be considered when interpreting results. First, although NDWA-UIC CUED investigators took extensive steps to collect a rich set of covariates and ensure their sample reflected city-specific DW labor force characteristics, we analyzed cross-sectional data from a non-random sample of informally employed DWers. Risk ratios may be underestimated as a result of the healthy worker survivor effect, in which both healthier and more exposed workers are more likely to remain in the active DW labor force and, thus, participate in the survey.[33] Moreover, pre-domestic work employment health status was not measured and may also lead to an underestimation of results (i.e., healthy worker hire effect). Second, although we excluded the only two exposure variables (sexual harassment, sexual assault) we assessed as severely underreported, measurement error of other self-reported hazards, which may vary by participant characteristics, could still bias results (i.e., overestimating the number of latent classes; incorrectly selecting the splitting variables for classification trees; underestimating exposure-outcome associations). Third, results may not be generalizable to contemporary informally employed US DWers. In our recent study using the same data,[12] we partially investigated this concern. We empirically examined—and observing similarities between—the individual, household, and employment characteristics of DWers in this unique 2011-2012 sample relative to those of DWers in the only other nationwide survey of formally and informally employed DWers, which lacked workplace hazards data and was conducted in 2020-2021, solely among Spanish-speaking DWers.[34]

Three lines of evidence lend credibility to our findings. First, the limited literature quantitatively examining exposure to single workplace hazards and health among DWers, in the US and internationally, documents cross-sectional associations of similar magnitude to those observed in this study using the single hazards approach. [7-11] For example, in a 2007 nationally representative sample of formally employed (i.e., agency-employed) home health aides, the odds of having a back injury were 4.64 (95% CI 1.6 to 14.0) times higher among aides who did versus did not report the need for additional ergonomic equipment, net of education, income, racialized group, and job training.[7] Second, a robust qualitative literature on the structure and conditions of DW describes similar patterns of exposures and related health concerns to those identified in our classification tree and latent class approaches.[35-38] For example, in 2007 Hondagneu-Sotelo [35] and in 2002 Romero [36] each reported how cleaning-related hazards cluster together among US DWers primarily responsible for housecleaning, often leading to back pain and injury. Finally, in addition to our recent study that developed the latent class approach further examined here,[12] the only other study we know of that quantitatively analyzed DWers’ joint exposures—which used multiple correspondence and clustering methods but did not examine health outcomes—identified three clusters of workplace abuse among a sample of Portuguese DWers.[39] More broadly, patterns of exposure to multiple hazards such as those observed in our study, and their association with multiple health outcomes, is akin to what other low-wage workers, in the US and internationally, experience at work.[1, 2, 40]

This study addresses an important gap in the literature by providing novel quantitative evidence regarding the work-related and general health of DWers experiencing distinct combinations of workplace hazards and showing the value of using diverse methods to quantify these joint exposure patterns. Taken together, our findings support the use of a latent class approach for identifying potential subgroups of workers unduly burdened and harmed, across multiple health metrics, by employer practices. More broadly, our results underscore the importance of conceptualizing and operationalizing measures that capture the range and patterns of DWers’ exposures. Future research on DWers should analyze joint patterns of exposure, examine the causal role of policy exclusions and employer practices in patterning DWers’ health (including collecting the contemporary, nationally representative, and longitudinal data needed to do so), and investigate both social inequities in health among DWers and also between DWers and more privileged workers.

## Data Availability

The 2011-2012 NDWA-UIC CUED survey data underlying this article cannot be shared publicly per the policies of NDWA to protect the privacy of individuals whose information was collected in these surveys. These terms of use are stipulated in the requirements of the authors' Data Use Agreement with the University of Illinois Chicago.

## ACKNOWLEDGMENTS

The authors acknowledge Paulina López González for her non-author contributions to interpreting the results of the latent class analysis for this manuscript, as well as her contributions (as a co-author) to our recently published manuscript in which we developed the latent class approach employed in this analysis. The authors acknowledge the National Domestic Workers Alliance (NDWA) and the University of Illinois Chicago Center for Urban Economic Development (UIC CUED) for providing the 2011-2012 NDWA-UIC CUED survey data. Nik Theodore thanks the Ford Foundation, Open Society Foundations, and Alexander Soros Foundation for funding the initial study that is the source of the 2011-2012 NDWA-UIC CUED survey data for this secondary analysis.

## CONTRIBUTORSHIP STATEMENT

Emily Wright initiated the study; led writing of the manuscript, conceptualizing the study design, acquiring the data, conducting the analyses, and interpreting the results; approved submission of the version to be published; and agrees to be accountable for all aspects of the work.

Jarvis T. Chen contributed to: conceptualizing the study, designing the analyses, interpreting the results, and preparing the manuscript; approved submission of the version to be published; and agrees to be accountable for all aspects of the work.

Jason Beckfield contributed to: conceptualizing the study, designing the analyses, interpreting the results, and preparing the manuscript; approved submission of the version to be published; and agrees to be accountable for all aspects of the work.

Nik Theodore led the original study from which the secondary 2011-2012 NDWA-UIC CUED data are derived; assisted with data acquisition; contributed to interpreting the results and preparing the manuscript; approved submission of the version to be published; and agrees to be accountable for all aspects of the work.

Nancy Krieger contributed to: conceptualizing the study, designing the analyses, interpreting the results, and preparing the manuscript; approved submission of the version to be published; and agrees to be accountable for all aspects of the work.

## ETHICAL APPROVAL

This study was determined to be Not Human Subjects Research (IRB21-0855) by the Harvard T.H. Chan School of Public Health Institutional Review Board.

