## Supplemental Methods for "Workplace hazards and health among informally employed domestic workers in 14 cities, United States, 2011-2012: using four approaches to characterize workers’ patterns of exposures"

Emily Wright

Jarvis T. Chen

Jason Beckfield

Nik Theodore

Nancy Krieger

**Affiliations:**

Harvard T.H. Chan School of Public Health, Department of Social and Behavioral Sciences, Boston, MA, USA (EW, JTC, NK)

Harvard University, Department of Sociology, Cambridge, MA, USA (JB)

University of Illinois Chicago, Department of Urban Planning and Policy, Chicago, IL, USA (NT)

**Correspondence to:** Emily Wright, Department of Social and Behavioral Sciences (Kresge 7^th^ floor), Harvard T.H. Chan School of Public Health, 677 Huntington Avenue, Boston, MA, USA 02115 (phone: 401-419-8629 |)

**Supplemental methods**

***Exclusion of sexual harassment and sexual assault variables***

We excluded the variables for sexual harassment and sexual assault from our analysis due to severe underreporting. Only 2.4% of workers in the NDWA-UIC CUED sample reported sexual harassment or assault in the last 12 months. In comparison, 27.6% of workers in an early 2010s sample of Oregon Medicaid homecare workers reported workplace sexual harassment in the last 12 months.[1] Studies of other US samples of low-wage [2] and predominantly women workers [3] from around the same time have also estimated a 12-month prevalence of sexual harassment of over 20%. Moreover, in a nationally representative sample of noninstitutionalized US adults from 2010 to 2012, 5.6% of women reported some type of sexual violence by a workplace-related perpetrator over the course of their lifetime—more than twice the 12-month prevalence among domestic workers (DWers) in our sample.[4]

Differences in the approach to measuring sexual harassment may partially explain its underreporting in the NDWA-UIC CUED sample, relative to other literature. NDWA-UIC CUED surveyors measured exposure to sexual harassment by asking participants if, in the last 12 months at any of their domestic work (DW) jobs, an employer or anyone in an employer’s home ever “Touched or looked at you, or behaved, in a sexually suggestive manner” or, separately, “Sexually assaulted you.” Both questions had response options for “yes” and “no.” The aforementioned studies reporting higher estimates [1-4] employed more behaviorally specific questions, often worded and with response options regarding the frequency of these experiences (e.g., “How many people have ever fondled, groped, grabbed, or touched you in a sexual way when you did not want it to happen?”). A 2020 expert review of workplace sexual harassment by the US Government Accountability Office reported that “Estimates of workplace sexual harassment based on surveys that provide respondents with examples of behaviors that may constitute sexual harassment were higher than other estimates based on surveys that ask directly about sexual harassment.”[5, p.12]

We conducted sensitivity analyses in which we included a composite variable for “any sexual harassment” (representing exposure to sexual harassment or sexual assault) in all phases of the analysis. As expected, results regarding the construction of the *a priori* composite, classification tree, and latent class exposure variables, as well as their association with the three health outcomes, were nearly identical (results available upon request).

Comparisons of the percent of workers reporting other exposures measured in the NDWA-UIC CUED sample to other literature did not raise similar concerns about underreporting. For example, a 2019 survey of New Jersey DWers that used many of the same items as the NDWA-UIC CUED survey reported similar percentages of workers being paid late (20%), paid less than agreed (12%), being paid with a bad check (7%), and being pressured to work more than one’s scheduled hours (16%) as were reported in our sample (respectively, 23.5%, 11.6%, 3.4%, and 18.8%).[6]

***Covariate selection***

The following covariates were included in all health outcome models: racialized group, citizenship and immigration status, gender, age, education (***individual covariates***), main DW occupation, live-in status, years worked as a DWer, hours worked last week for one’s main DW employer, number of DW employers, had a contract with any of their DW employers in the last 12 months (***occupational covariates***), number of people relying on the participant’s financial support, income earner status, and household economic insecurity (***household covariates***). We constructed a composite categorical variable reflecting how many forms of economic insecurity a participant’s household experienced (2=paid an essential bill late in the last month and paid rent or mortgage late in the last 12 months [n=634]; 1=one of these [n=502]; 0=neither [n=904]). We did not include any city- or state-level covariates because all health outcome models included city fixed effects.

Covariate selection was primarily guided by our use of ecosocial theory.[7-9] We conceptualized DWers as an occupational-social group shaped by multi-level non-nested dynamic power relations involving racism, sexism, immigration policies, and economic deprivation. Based on this, we expected that covariates capturing participants’ membership in social groups defined by these types of injustices (e.g., racialized group, citizenship and immigration status, gender, age, education, number of people relying on the participant’s financial support, income earner status, household economic insecurity) would be associated with both their workplace exposures and health. We also expected that occupational characteristics such as main DW occupation, live-in status, hours worked, number of DW employers, and contract status would be associated with both DWers’ workplace exposures (e.g., housecleaners are more likely to be responsible for hazardous cleaning-related tasks than adult care workers; DWers working for more than one employer may be more likely to experience a given hazard at one or more of their multiple employers than DWers with only one employer) and health (e.g., workers with a written contract may be more likely to receive health-related benefits such as paid sick leave). Finally, we included the number of years a participant had worked as a DWer in the US to partially address the healthy worker survivor effect, whereby the healthiest workers disproportionately remain employed in hazardous jobs over time.[10] Although the NDWA-UIC CUED survey also collected data on health insurance status, we did not include this covariate in analytic models since we hypothesized it may partially mediate the associations of interest. Data on health insurance status is included in descriptive tables (**Table 1**).

Covariate selection was also informed by prior research on DWers and their health. Robust qualitative social science literature on DW in the US illustrates how racialized group membership, citizenship and immigration status, main DW occupation, age, live-in status, educational attainment, and social class pattern workplace exposures.[11-14] The limited quantitative literature on the working conditions and health of informally employed DWers in the US and internationally also documents that exposure to multiple workplace hazards, as well as health, may be patterned by DWers’ individual and occupational characteristics.[15-20]

Finally, covariate selection was informed by observed variation in DWer exposures and health by covariates. Descriptive analyses of the percent of DWers exposed to each of the hazards suggested that racialized group, gender, education, citizenship and immigration status, main DW occupation, live-in status, household economic insecurity, and, to some extent, age, contract status, and years worked as a DWer in the US were associated with variation in exposures (**Figure S1**).

We examined for model fitting problems related to collinearity and model complexity by including the city fixed effects and selected covariates in the health outcome models in five steps. First, we included only the exposure of interest (“M1”). Second, we added the city fixed effects to the models (“M2”). Then, we added all aforementioned individual-level covariates to the models (“M3”). Next, we added all occupational covariates to the models (“M4”). Finally, we included household covariates (“M5”). We inspected model results (**Figure S5**) from each step for changes to the point estimates and widening of the confidence intervals, above and beyond what would be expected by covariate control, as these are some of the implications of predictor collinearity and model overfitting. We also examined collinearity using the generalized variance inflation factor (VIF), which assesses the degree to which the variance of each parameter estimate is inflated due to the collinearity of the predictor variable for that parameter with all other variables in the model. VIF ranges from one to infinity, with larger values indicating greater inflation. For all three health outcomes, the VIF for all covariates was <3, and the vast majority were <2, indicating minimal to acceptable levels of collinearity.[21, 22]

In the fully-adjusted models (M5), we conducted sensitivity analyses regarding unmeasured confounding using the E-value.[23] E-values represent the minimum strength of the association, on the risk ratio scale, that unmeasured confounder(s) would need to have with both the exposure and the outcome to fully explain away the estimated exposure-outcome association, conditional on all covariates included in the fully-adjusted health outcome models.

***Missing data***

We used multiply imputed data on the health outcomes and covariates in the latent class analysis and health outcome models using all four exposure categorization approaches. Multiple imputation models rely on the Missing-At-Random (MAR) assumption, meaning that the probability that a variable is missing can be related to the observed data, but not to the missing data itself. Although the MAR assumption cannot be empirically evaluated, this assumption seemed reasonable because a) marginal missingness was low (0.2-1.2% of participants were missing data on any of the workplace hazards; 0.0-6.9% on the selected covariates; and 0.3-1.9% on the health outcomes), b) missingness on most variables was not predicted by other observed variables, c) missingness on other analytic variables (e.g., citizenship and immigration status [6.9% missing]) was related to other observed variables (e.g., place of birth, age, number of years lived in the US), and d) evidence from simulation studies suggests that if the missing data rate does not exceed 25% and if the correlation between the unobserved covariate and the dependent variable is 0.4, omitting an unobserved variable from the imputation model has a negligible effect.[24] We also conducted sensitivity analyses in which we used complete cases only in the health outcome modeling series. Results were robust to the use of these different missing variable methods, including regarding the direction, magnitude, and significance of point estimates related to the workplace hazard exposure variables (results available upon request).

We defined the predictor matrix for the final multiple imputation model using the quickpred() function—using default values for the minimum target variable-predicator variable correlation threshold (0.1) and the minimum proportion of usable cases (zero)—and the following variables:[25]

a. ***Individual:*** gender, racialized group, educational attainment, citizenship and immigration status, marital status, birth place, city, work-related back injury in the last 12 months, work-related illness in the last 12 months, self-rated health;
b. ***Household:*** paid rent or mortgage late at any time last 12 months, paid any essential bills late last month, saved any money for the future in the last month, currently spent over half of their income on rent or mortgage, income earner status, parent to one or more children under 18 years of age living in the US, had no food to eat at home in the last month, health insurance status, number of family members supported on participant’s income, whether the participant supported family abroad by sending money, years lived in the US; and
c. ***Employment and occupational:*** live-in status, main DW occupation, problems with working conditions in the last 12 months, whether a participant worked another job besides domestic work in the last week, years worked with their main DW employer, hours worked last week with their main DW employer, whether they had a contract with any DW employer in the last 12 months, whether they perceived any of their current DW jobs to be dangerous or hazardous, years worked as a DWer, number of DW employers in the past 30 days.

For the few variables with response options for “don’t know”—including whether DWers had an employment contract, perceived any of their DW jobs as dangerous or hazardous, had paid their rent or mortgage late in the last 12 months, and currently spent more than half of their income on rent or mortgage—we grouped the few participants who responded “don’t know” (<2% of the total sample of each variable) with those missing for imputation.

***Rationale for selection of the a priori hazards for composite exposures***

Work-related back injury. We selected “heavy lifting,” “climbing to clean,” and “working long hours with no breaks” as the three hazards to be included in the composite exposure measure for work-related back injury. We selected “heavy lifting” because of the strain it places on muscles, tendons, and ligaments of the back, especially when done in awkward postures, which may often be the case for DWers lifting a) patients in patients’ homes without assistive equipment and/or b) furniture and other large objects to clean (under) them. A large occupational health literature documents associations between exposure to heavy lifting and risk of back injury.[26, 27] We considered both “climbing to clean” and “being required to work on one’s knees” given documented associations between trunk flexion and other demanding postures and the risk of back injury.[26] We selected the former because we expected the risk of back injury might be higher among individuals who climbed to clean, rather than knelt, due to the risk of forceful landing or fall when climbing down from cleaning. Finally, we selected “working long hours with no breaks” because we expected this variable—especially in combination with the other two selected variables—might capture cumulative back strain caused by repetitive activity and without adequate time for recovery in between tasks.[28]

Work-related illness. We selected “caring for someone with a contagious illness” and “working long hours with no breaks” to be included in the composite exposure measure for work-related illness. We selected “caring for someone with a contagious illness” because this was the only variable that captured DWers’ risk of exposure to contagious illness on the job.[29] We selected “working long hours with no breaks” because we expected this variable—especially in combination with the first variable—would help further capture DWers’ risk of exposure to contagious illness on the job, via prolonged contact with individuals with a contagious illness. We also expected that DWers who worked long hours without breaks might be at higher risk of contracting a contagious illness given exposure on the job, through pathways related to immunological function.[30-32]

Fair-to-poor self-rated health. We selected “working long hours with no breaks,” “working with toxic cleaning supplies,” and “being threatened” to be included in the composite exposure measure for fair-to-poor self-rated health. We selected “working long hours with no breaks” because this may contribute to fatigue, sleep deprivation (especially for live-in workers), risk of on-the-job injury and illness, work-related stress and other threats to general health. We selected “working with toxic cleaning supplies” since the use of such products may be linked to skin and eye irritation, trouble breathing, and other respiratory symptoms and conditions.[33-35] Finally, although we expected several hazards (e.g., “verbally abused or yelled at”; “called insulting names or racial slurs”; “used immigration status to threaten”; “threatened”) may be linked to self-rated health via pathways related to discrimination and other forms of socially inflicted trauma,[36] we selected “threatened.” This is because we expected the stress induced by the threatening of specific harm(s) (e.g., loss of employment, physical harm, calling the police) might meaningfully and quickly undermine self-rated health.

***Classification trees***

We fit classification trees using the rpart() function in R.[37] Each classification tree model included one of the three binary health outcomes and all 19 binary workplace hazard variables. Models were fit using observed (i.e., non-imputed) data on these variables. Any DWer with observed data for the health outcome and at least one workplace hazard participated in the classification tree modeling. This means that the Gini index (described below), as well as the probability that individuals belong to a particular leaf of the tree, is calculated only for the individuals not missing data on a particular hazard being considered by the model. For individuals missing data on a variable that ultimately determines a split in the tree, rpart() uses observed information on other variables, called surrogates, to assign them to a given leaf.

We used the Gini index of impurity as the splitting criterion for our classification trees—to recursively determine the which hazard should be used to “best” group individuals according to their risk of the health outcome. The Gini measure of impurity ranges from zero to 0.5. Zero indicates maximum purity, meaning that all individuals assigned to one leaf of the tree according to their exposures have the same health outcome. A Gini index value of 0.5 indicates maximum impurity, meaning that only half of the individuals assigned to one leaf reported the same health outcome. In applications with binary outcomes such as our study, the Gini index and the Information index—the alternative impurity measure available in rpart()—will almost always choose the same variable splits.

Procedurally, classification trees calculate the Gini index for all possible splits and use the split that maximizes the reduction in impurity (i.e., increases the homogeneity of the outcome variable within the leaves of the tree). This process continues until no more useful splits can be found or, alternatively, a pre-set minimum number of individuals is observed in the leaves of the tree. Splits are no longer useful when they fail to decrease the lack of fit of the model to the data by a given amount. This amount is referred to as the “complexity parameter,” which can be thought of as the “cost” of adding another variable to the classification model. We used the default rpart() values for the complexity parameter (cp=0.01), the minimum number of individuals that must be assigned to a branch for the model to attempt to compute a further split of it (n=20), and the minimum number of observations that must exist in each terminal node of the tree (n=7).

Because the classification tree resulting during this initial phase of modeling is often unnecessarily complex—despite the inclusion of an initial complexity parameter—a second analytic phase is used to determine how to “prune” the tree, maintaining only the most useful branches and leaves. This second phase consists, first, of randomly splitting the dataset into *k* sub-datasets, fitting (also known as “training”) a classification tree on all but one of the sub-datasets, and then evaluating the performance of the model in predicting the health outcome on the remaining sub-dataset (also known as “testing”). This method is referred to as cross-validation. We conducted 10 cross-validations, the rpart() default. Based on the results of the cross-validation, we used the 1-standard error rule to determine how to prune the trees (i.e., how many branches and leaves to retain).

We plotted the classification tree results using rpart.plot() in R.[38] For each terminal leaf of the tree, these plots show a) the predicted health outcome for members of that group, b) the predicted probability of experiencing the health outcome of interest, and c) the percentage of observations assigned to that leaf of the tree.

We chose to use classification trees rather than random forest models. Random forests is another supervised learning method that randomly builds an ensemble of classification trees and then merges them together to form a more stable and accurate solution. Despite this advantage, visualizing and interpreting the results of a random forest model is much less intuitive. Because the objective of our paper was to compare a suite of exposure categorization methods, rather than rely on one for the most accurate and stable estimates, we prioritized interpretability and ease of communication of the results to the reader and used classification trees.

***Latent class analysis***

We conducted our latent class analysis in three phases: class enumeration (Phase 1), model building (Phase 2), and estimation of associations between latent class membership and the three health outcomes (Phase 3). The methods for Phase 1 and Phase 2 were developed and are presented in our recent manuscript—see Wright et al for more details on and results from this approach.[39] Briefly, in **Phase 1** we determined the number of latent classes to retain in the model, without covariates. We initially fit Phase 1 models with all 19 workplace hazard variables. We monitored these models for violations of the LCA conditional independence assumption, which states that membership in the latent classes explains all associations between the indicators, and can arise due to redundancies between variables. Such violations, by causing fit indices to increase, can lead to the selection of more classes than is truly necessary.[40] Because we observed conditional independence violations—as expected, between hazards of the same “type” (i.e., economic, social, or occupational)—in all initial models, we combined redundant indicators (e.g., “hurt or attacked” and “pushed or shoved”) into composite indicators (e.g., “any physical attack”) and estimated Phase 1 models with this reduced set of 10 indicators before deciding how many latent classes to retain.[41]

In **Phase 2** (model building), we monitored for other sources of conditional independence violations and added model features to address them as needed. Conditional independence violations can also arise due to variation in the statistical associations between indicators varies across covariates, beyond what is explained by latent class membership. We tested for this by adding each of a set of selected covariates (main occupation, live-in status, citizenship and immigration status, racialized group, educational attainment, age, and household economic insecurity) to the final Phase 1 model.[42] We successively added all covariates that demonstrated significant associations with indicators (main occupation, live-in status, citizenship and immigration status, and racialized group) to the model, as predictors of latent class membership and the relevant indicators. Then, for any pairs of indicators with residual associations that continued to violate conditional independence, we successively added “direct effects” between them. These direct effects allow pairs of indicators to correlate freely within classes, relaxing the conditional independence assumption.[41] Again, we expected, and ultimately observed, such residual associations would occur between pairs of hazard variables of the same “type” (i.e., occupational, social, or economic hazards). We successively added direct effects between pairs of indicators with the largest remaining residual associations until no significant residual associations remained, selecting the four-class model in which only two significantly elevated, but qualitatively small, residual associations remained as the final model. Phase 1 and 2 latent class models used full information maximum likelihood to handle the minimal missing data on the workplace hazard indicators under the MAR assumption.[43]

After building the final LCA model, in **Phase 3** we examined associations between latent class membership and three health outcomes, using the multiply imputed covariate and health outcome data. Specifically, we used the maximum likelihood bias-adjusted three-step approach with modal class assignment to estimate these associations.[42, 44, 45] This approach involved fitting the final LCA model from Phase 2, obtaining the classifications from it, and fitting city fixed effects log Poisson regression models—one for each health outcome of interest—to estimate the associations, while taking into account the uncertainty in latent class assignment. We obtained estimated risk ratios and cluster robust 95% confidence intervals of each health outcome from these models.
