## Supplementary material for "Workplace hazards and health among informally employed domestic workers in 14 cities, United States, 2011-2012: using four approaches to characterize workers’ patterns of exposures": Table S1

| **Table S1. Estimated item response probabilities and robust standard errors from final four-class latent class analysis model of domestic workers’ patterns of workplace hazard exposure: NDWA-UIC CUED data, 14 cities, United States, 2011-2012 (N = 2,086).** | | | | | | | | |
| --- | --- | --- | --- | --- | --- | --- | --- | --- |
| **Workplace hazard** | **Latent class** | | | | | | | |
|  | **Class 1:**  **Low hazard domestic work** | | **Class 2:**  **Demanding care work** | | **Class 3:**  **Strenuous cleaning work** | | **Class 4:**  **Hazardous domestic work** | |
|  | **Probability** | **Robust standard error** | **Probability** | **Robust standard error** | **Probability** | **Robust standard error** | **Probability** | **Robust standard error** |
| Any pay violation | 0.116 | 0.017 | 0.421 | 0.079 | 0.298 | 0.046 | 0.674 | 0.060 |
| Any verbal abuse | 0.045 | 0.015 | 0.227 | 0.038 | 0.116 | 0.032 | 0.523 | 0.079 |
| Any physical attack | 0.003 | 0.003 | 0.048 | 0.021 | 0.014 | 0.007 | 0.095 | 0.028 |
| Any immigration status threat | 0.000 | 0.000 | 0.022 | 0.012 | 0.015 | 0.011 | 0.164 | 0.036 |
| Cared for someone with contagious illness | 0.007 | 0.009 | 0.381 | 0.133 | 0.030 | 0.017 | 0.211 | 0.045 |
| Did heavy lifting or other strenuous activities | 0.056 | 0.031 | 0.452 | 0.061 | 0.498 | 0.091 | 0.943 | 0.034 |
| Climbed to clean | 0.006 | 0.019 | 0.008 | 0.018 | 0.869 | 0.096 | 0.946 | 0.026 |
| Required to work on knees | 0.050 | 0.017 | 0.286 | 0.058 | 0.548 | 0.058 | 0.766 | 0.035 |
| Worked with toxic cleaning supplies | 0.182 | 0.022 | 0.243 | 0.084 | 0.761 | 0.038 | 0.863 | 0.027 |
| Worked long hours without breaks | 0.062 | 0.022 | 0.574 | 0.104 | 0.311 | 0.060 | 0.888 | 0.084 |
| Abbreviations: NDWA-UIC CUED = National Domestic Workers Alliance and University of Illinois Chicago Center for Urban Economic Development | | | | | | | | |
