## Supplementary material for "Workplace hazards and health among informally employed domestic workers in 14 cities, United States, 2011-2012: using four approaches to characterize workers’ patterns of exposures": Table S2

| **Table S2. Estimated risk ratios and 95% confidence intervals of work-related back injury, work-related illness, and fair-to-poor self-rated health from fully-adjusted city fixed effects models using *a priori* composite exposure, classification tree, and latent class approaches to defining exposure: NDWA-UIC CUED data, 14 cities, United States, 2011-2012 (N = 2,086).** | | | | | | | | | |
| --- | --- | --- | --- | --- | --- | --- | --- | --- | --- |
| **Variable** | **Work-related back injury** | | | **Work-related illness** | | | **Fair-to-poor self-rated health** | | |
|  | ***A priori* composite exposure** | **Classification tree** | **Latent class analysis** | ***A priori* composite exposure** | **Classification tree** | **Latent class analysis** | ***A priori* composite exposure** | **Classification tree** | **Latent class analysis** |
|  | **RR (95% CI)** | **RR (95% CI)** | **RR (95% CI)** | **RR (95% CI)** | **RR (95% CI)** | **RR (95% CI)** | **RR (95% CI)** | **RR (95% CI)** | **RR (95% CI)** |
| **Intercept (baseline risk)** | 0.0 (0.0 to 0.1) | 0.1 (0.0 to 0.1) | 0.0 (0.0 to 0.0) | 0.1 (0.1 to 0.3) | 0.1 (0.1 to 0.3) | 0.1 (0.0 to 0.1) | 0.1 (0.1 to 0.2) | 0.2 (0.1 to 0.2) | 0.1 (0.1 to 0.2) |
| **Exposure** |  |  |  |  |  |  |  |  |  |
| AP: None/CT: No hazard #1/LCA: Class 1^a^ | Ref | Ref | Ref | Ref | Ref | Ref | Ref | Ref | Ref |
| AP: Some/CT: Hazard #1/LCA: Class 2^b^ | 3.2 (2.5 to 4.1) | 3.0 (2.3 to 3.9) | 3.1 (2.0 to 4.9) | 1.9 (1.6 to 2.3) | 1.7 (1.4 to 2.0) | 2.2 (1.6 to 3.0) | 1.3 (1.1 to 1.5) | 1.4 (1.2 to 1.7) | 1.0 (0.7 to 1.5) |
| AP: All/CT: Hazard #1 and #2/LCA: Class 3^c^ | 6.5 (4.8 to 8.7) | 4.4 (3.6 to 5.3) | 3.7 (2.6 to 5.2) | 2.9 (2.4 to 3.6) | 2.6 (2.2 to 3.2) | 2.1 (1.6 to 2.7) | 1.8 (1.4 to 2.3) | 1.9 (1.4 to 2.4) | 1.5 (1.2 to 1.8) |
| LCA: Class 4^d^ | Not applicable | Not applicable | 6.6 (4.6 to 9.4) | Not applicable | Not applicable | 2.7 (2.0 to 3.5) | Not applicable | Not applicable | 1.6 (1.3 to 2.0) |
| **Racialized group** |  |  |  |  |  |  |  |  |  |
| White | Ref | Ref | Ref | Ref | Ref | Ref | Ref | Ref | Ref |
| Latina/o | 1.5 (1.1 to 2.0) | 1.4 (1.0 to 2.0) | 1.6 (1.2 to 2.1) | 1.1 (0.8 to 1.3) | 1.0 (0.8 to 1.3) | 1.0 (0.8 to 1.3) | 1.4 (1.2 to 1.7) | 1.4 (1.2 to 1.7) | 1.5 (1.1 to 1.9) |
| Black | 1.2 (0.7 to 1.9) | 1.1 (0.6 to 1.9) | 1.2 (0.8 to 1.9) | 0.6 (0.4 to 1.0) | 0.6 (0.3 to 1.1) | 0.6 (0.4 to 0.9) | 0.7 (0.4 to 1.1) | 0.7 (0.4 to 1.1) | 0.7 (0.5 to 1.1) |
| “Asian or other” | 0.9 (0.6 to 1.4) | 0.8 (0.5 to 1.3) | 0.9 (0.6 to 1.4) | 0.7 (0.5 to 0.9) | 0.6 (0.5 to 0.9) | 0.6 (0.4 to 0.9) | 1.1 (0.9 to 1.4) | 1.2 (1.0 to 1.5) | 1.2 (0.9 to 1.7) |
| **Citizenship and immigration status** |  |  |  |  |  |  |  |  |  |
| US citizen | Ref | Ref | Ref | Ref | Ref | Ref | Ref | Ref | Ref |
| Documented immigrant | 1.1 (0.9 to 1.5) | 1.2 (0.9 to 1.7) | 1.1 (0.8 to 1.4) | 0.9 (0.8 to 1.2) | 0.9 (0.8 to 1.2) | 0.9 (0.7 to 1.1) | 1.1 (0.9 to 1.3) | 1.1 (1.0 to 1.3) | 1.1 (0.9 to 1.4) |
| Undocumented immigrant | 1.1 (0.8 to 1.6) | 1.2 (0.9 to 1.6) | 1.1 (0.9 to 1.5) | 1.3 (1.0 to 1.5) | 1.2 (1.0 to 1.5) | 1.2 (0.9 to 1.5) | 1.2 (0.9 to 1.4) | 1.1 (0.9 to 1.4) | 1.1 (0.9 to 1.4) |
| **Gender** |  |  |  |  |  |  |  |  |  |
| Women | Ref | Ref | Ref | Ref | Ref | Ref | Ref | Ref | Ref |
| Men | 1.3 (1.0 to 1.6) | 1.4 (1.0 to 1.8) | 1.4 (0.9 to 2.1) | 0.8 (0.5 to 1.4) | 0.8 (0.5 to 1.4) | 0.7 (0.4 to 1.3) | 0.7 (0.4 to 1.3) | 0.7 (0.4 to 1.3) | 0.7 (0.4 to 1.3) |
| **Age** |  |  |  |  |  |  |  |  |  |
| <25 | Ref | Ref | Ref | Ref | Ref | Ref | Ref | Ref | Ref |
| 25-44 | 1.0 (0.7 to 1.5) | 1.0 (0.7 to 1.4) | 1.0 (0.6 to 1.6) | 1.1 (0.8 to 1.6) | 1.1 (0.8 to 1.5) | 1.1 (0.8 to 1.5) | 1.2 (1.0 to 1.5) | 1.2 (0.9 to 1.4) | 1.2 (0.8 to 1.8) |
| 45-64 | 1.3 (0.9 to 1.9) | 1.3 (0.9 to 1.8) | 1.3 (0.8 to 2.1) | 1.0 (0.7 to 1.4) | 1.0 (0.7 to 1.4) | 1.0 (0.7 to 1.4) | 1.6 (1.2 to 2.0) | 1.6 (1.2 to 2.1) | 1.6 (1.0 to 2.4) |
| ≥65 | 1.4 (0.8 to 2.3) | 1.2 (0.7 to 2.0) | 1.3 (0.6 to 2.5) | 0.9 (0.5 to 1.6) | 0.9 (0.5 to 1.5) | 0.9 (0.5 to 1.6) | 1.7 (1.2 to 2.6) | 1.7 (1.2 to 2.6) | 1.7 (1.0 to 3.0) |
| **Formal educational attainment** |  |  |  |  |  |  |  |  |  |
| Less than 12 years | Ref | Ref | Ref | Ref | Ref | Ref | Ref | Ref | Ref |
| ≥HSD (or equivalent) and <Bachelor’s degree | 1.1 (0.9 to 1.3) | 1.2 (1.0 to 1.4) | 1.1 (0.9 to 1.3) | 1.2 (0.9 to 1.4) | 1.2 (0.9 to 1.4) | 1.1 (0.9 to 1.3) | 0.8 (0.7 to 0.9) | 0.8 (0.7 to 0.9) | 0.8 (0.7 to 0.9) |
| Bachelor’s degree or higher | 1.3 (1.0 to 1.7) | 1.3 (1.0 to 1.8) | 1.2 (0.8 to 1.6) | 1.2 (1.0 to 1.5) | 1.2 (1.0 to 1.5) | 1.1 (0.8 to 1.5) | 0.6 (0.5 to 0.8) | 0.6 (0.5 to 0.8) | 0.6 (0.4 to 0.8) |
| **Main occupation** |  |  |  |  |  |  |  |  |  |
| Housecleaning | Ref | Ref | Ref | Ref | Ref | Ref | Ref | Ref | Ref |
| Child care | 1.0 (0.8 to 1.2) | 0.9 (0.7 to 1.1) | 1.0 (0.8 to 1.3) | 1.2 (0.9 to 1.4) | 1.2 (0.9 to 1.5) | 1.4 (1.1 to 1.8) | 0.9 (0.8 to 1.1) | 1.0 (0.8 to 1.1) | 1.0 (0.8 to 1.3) |
| Adult care | 1.3 (1.0 to 1.7) | 1.1 (0.9 to 1.4) | 1.3 (1.0 to 1.7) | 1.0 (0.8 to 1.3) | 1.0 (0.8 to 1.3) | 1.0 (0.8 to 1.4) | 1.3 (1.1 to 1.6) | 1.3 (1.1 to 1.6) | 1.4 (1.1 to 1.7) |
| **Live-in** |  |  |  |  |  |  |  |  |  |
| No | Ref | Ref | Ref | Ref | Ref | Ref | Ref | Ref | Ref |
| Yes | 1.0 (0.7 to 1.3) | 1.0 (0.8 to 1.4) | 0.9 (0.7 to 1.3) | 1.0 (0.8 to 1.3) | 1.0 (0.8 to 1.3) | 1.0 (0.8 to 1.3) | 1.1 (0.9 to 1.4) | 1.1 (0.9 to 1.4) | 1.1 (0.8 to 1.4) |
| **Years as DWer in the US** |  |  |  |  |  |  |  |  |  |
| <5 | Ref | Ref | Ref | Ref | Ref | Ref | Ref | Ref | Ref |
| 5-9 | 1.0 (0.8 to 1.3) | 1.0 (0.8 to 1.3) | 1.0 (0.8 to 1.3) | 1.1 (0.9 to 1.3) | 1.1 (0.9 to 1.4) | 1.1 (0.9 to 1.4) | 0.9 (0.8 to 1.1) | 0.9 (0.8 to 1.1) | 0.9 (0.8 to 1.1) |
| 10-19 | 1.2 (1.0 to 1.6) | 1.2 (1.0 to 1.5) | 1.3 (1.0 to 1.6) | 1.0 (0.8 to 1.3) | 1.0 (0.8 to 1.3) | 1.1 (0.9 to 1.3) | 1.0 (0.9 to 1.2) | 1.0 (0.8 to 1.1) | 1.0 (0.8 to 1.2) |
| ≥20 | 1.2 (0.8 to 1.7) | 1.2 (0.9 to 1.7) | 1.1 (0.8 to 1.6) | 0.9 (0.7 to 1.2) | 0.9 (0.7 to 1.2) | 0.9 (0.6 to 1.3) | 1.1 (0.9 to 1.3) | 1.0 (0.8 to 1.2) | 1.0 (0.8 to 1.3) |
| **Working hours, main DW employer, last week** |  |  |  |  |  |  |  |  |  |
| <8 | Ref | Ref | Ref | Ref | Ref | Ref | Ref | Ref | Ref |
| 8-19 | 0.7 (0.6 to 0.9) | 0.7 (0.6 to 0.8) | 0.7 (0.6 to 1.0) | 1.0 (0.8 to 1.2) | 1.0 (0.8 to 1.3) | 1.1 (0.8 to 1.5) | 0.9 (0.8 to 1.1) | 0.9 (0.8 to 1.0) | 0.9 (0.7 to 1.2) |
| 20-39 | 0.9 (0.7 to 1.1) | 0.9 (0.7 to 1.1) | 0.9 (0.7 to 1.3) | 1.2 (0.8 to 1.7) | 1.2 (0.9 to 1.7) | 1.3 (1.0 to 1.8) | 0.9 (0.7 to 1.1) | 0.9 (0.7 to 1.0) | 0.9 (0.7 to 1.2) |
| ≥40 | 0.9 (0.7 to 1.2) | 0.9 (0.7 to 1.1) | 1.0 (0.7 to 1.4) | 1.2 (0.9 to 1.6) | 1.2 (0.9 to 1.6) | 1.4 (1.0 to 2.1) | 0.8 (0.7 to 1.0) | 0.8 (0.7 to 1.0) | 0.8 (0.6 to 1.2) |
| **Number of DW employers, last mo.** |  |  |  |  |  |  |  |  |  |
| 0-1 | Ref | Ref | Ref | Ref | Ref | Ref | Ref | Ref | Ref |
| 2 | 1.2 (1.1 to 1.4) | 1.2 (1.0 to 1.5) | 1.2 (1.0 to 1.6) | 1.0 (0.8 to 1.2) | 1.0 (0.8 to 1.2) | 1.0 (0.8 to 1.3) | 1.2 (1.0 to 1.4) | 1.1 (0.9 to 1.4) | 1.1 (0.9 to 1.4) |
| ≥3 | 1.1 (0.9 to 1.3) | 1.2 (1.0 to 1.5) | 1.1 (0.9 to 1.5) | 1.0 (0.8 to 1.3) | 1.0 (0.8 to 1.3) | 1.1 (0.8 to 1.4) | 1.1 (0.9 to 1.3) | 1.1 (0.9 to 1.2) | 1.1 (0.8 to 1.3) |
| **Contract, any DW employer, last 12 mo.** |  |  |  |  |  |  |  |  |  |
| No | Ref | Ref | Ref | Ref | Ref | Ref | Ref | Ref | Ref |
| Yes | 1.1 (1.0 to 1.3) | 1.0 (0.9 to 1.2) | 1.1 (0.9 to 1.4) | 0.9 (0.9 to 1.0) | 1.0 (0.9 to 1.1) | 1.0 (0.8 to 1.1) | 0.9 (0.8 to 1.0) | 0.9 (0.8 to 1.0) | 0.9 (0.8 to 1.0) |
| **Income earner status** |  |  |  |  |  |  |  |  |  |
| Joint earner | Ref | Ref | Ref | Ref | Ref | Ref | Ref | Ref | Ref |
| Main earner | 1.1 (0.9 to 1.4) | 1.2 (1.0 to 1.5) | 0.9 (0.7 to 1.1) | 0.9 (0.7 to 1.0) | 0.9 (0.7 to 1.1) | 1.1 (0.9 to 1.4) | 1.0 (0.9 to 1.1) | 1.0 (0.9 to 1.1) | 1.0 (0.8 to 1.3) |
| Sole earner | 1.0 (0.9 to 1.1) | 1.1 (1.0 to 1.2) | 0.9 (0.7 to 1.1) | 1.0 (0.8 to 1.2) | 1.0 (0.8 to 1.2) | 1.1 (0.9 to 1.4) | 1.2 (1.1 to 1.2) | 1.2 (1.1 to 1.2) | 1.2 (0.9 to 1.5) |
| **Number of people relying on financial support** |  |  |  |  |  |  |  |  |  |
| 0-1 | Ref | Ref | Ref | Ref | Ref | Ref | Ref | Ref | Ref |
| 2 | 1.4 (1.1 to 1.6) | 1.4 (1.2 to 1.7) | 1.3 (1.1 to 1.7) | 1.0 (0.8 to 1.2) | 1.0 (0.8 to 1.2) | 1.0 (0.8 to 1.2) | 1.2 (1.1 to 1.4) | 1.2 (1.0 to 1.3) | 1.2 (0.9 to 1.4) |
| ≥3 | 1.1 (0.9 to 1.4) | 1.2 (1.0 to 1.4) | 1.1 (0.9 to 1.4) | 1.0 (0.8 to 1.2) | 1.0 (0.8 to 1.2) | 0.9 (0.8 to 1.2) | 1.3 (1.1 to 1.5) | 1.2 (1.0 to 1.4) | 1.2 (1.0 to 1.5) |
| **Household economic insecurity** |  |  |  |  |  |  |  |  |  |
| Neither | Ref | Ref | Ref | Ref | Ref | Ref | Ref | Ref | Ref |
| Paid rent/mortgage or essential bill late | 1.2 (1.0 to 1.5) | 1.2 (1.0 to 1.5) | 1.2 (0.9 to 1.5) | 1.2 (1.0 to 1.4) | 1.1 (1.0 to 1.4) | 1.1 (0.9 to 1.4) | 1.3 (1.1 to 1.7) | 1.3 (1.0 to 1.6) | 1.3 (1.1 to 1.6) |
| Paid rent/mortgage and essential bill late | 1.3 (1.2 to 1.6) | 1.5 (1.3 to 1.8) | 1.3 (1.0 to 1.6) | 1.3 (1.1 to 1.4) | 1.3 (1.1 to 1.4) | 1.2 (1.0 to 1.4) | 1.6 (1.4 to 1.9) | 1.6 (1.3 to 1.9) | 1.6 (1.3 to 1.9) |
| Abbreviations: AP = *a priori* composite exposure; CI = confidence interval; CT = classification tree; DW = domestic work; DWer = domestic worker; HSD = high school diploma; LCA = latent class analysis; mo = month(s); NDWA-UIC CUED = National Domestic Workers Alliance and University of Illinois Chicago Center for Urban Economic Development; RR = risk ratio; US = United States. All confidence intervals are based on cluster robust standard errors. All results are from fully-adjusted models (M5) that include individual, occupational, and household covariates, as well as city fixed effects. City fixed effects estimates are not reported here, as required by the terms of the authors’ NDWA-UIC CUED data use agreement, which states that “At no point will the Recipient publish a direct comparison between cities included in the dataset or city-specific data.”  ^a^ “AP: None” refers to “Did no heavy lifting and no climbing to clean and did not work long hours with no break,” “No contagious illness care and did not work long hours with no breaks,” and “Did not work long hours with no breaks and did not work with toxic cleaning supplies and was not threatened” for the outcomes, respectively, of work-related back injury, work-related illness, and fair-to-poor self-rated health. “CT: No hazard #1” refers to “Did no heavy lifting,” “Did not work long hours with no breaks,” and “Did not climb to clean” for the outcomes, respectively, of work-related back injury, work-related illness, and fair-to-poor self-rated health. “LCA: Class 1” refers to the “Low hazard domestic work” class, for all outcomes.  ^b^ “AP: Some” refers to “Did heavy lifting or climbed to clean or worked long hours with no break,” “Did contagious illness care or worked long hours with no breaks,” and “Worked long hours with no breaks or worked with toxic cleaning supplies or was threatened” for the outcomes, respectively, of work-related back injury, work-related illness, and fair-to-poor self-rated health. “CT: Hazard #1” refers to “Did heavy lifting,” “Worked long hours with no breaks,” and “Climbed to clean” for the outcomes, respectively, of work-related back injury, work-related illness, and fair-to-poor self-rated health. “LCA: Class 2” refers to the “Demanding care work” class, for all outcomes.  ^c^ “AP: All” refers to “Did heavy lifting and climbed to clean and worked long hours with no break,” “Did contagious illness care and worked long hours with no breaks,” and “Worked long hours with no breaks and worked with toxic cleaning supplies and was threatened” for the outcomes, respectively, of work-related back injury, work-related illness, and fair-to-poor self-rated health. “CT: Hazard #1 and #2” refers to “Did heavy lifting and was verbally abused,” “Worked long hours with no breaks and did contagious illness care,” and “Climbed to clean and called insulting names or racial slurs” for the outcomes, respectively, of work-related back injury, work-related illness, and fair-to-poor self-rated health. “LCA: Class 3” refers to the “Strenuous cleaning work” class, for all outcomes.  ^d^ “LCA: Class 4” refers to the “Hazardous domestic work” class, for all outcomes. | | | | | | | | | |
