## Supplementary material for "Workplace hazards and health among informally employed domestic workers in 14 cities, United States, 2011-2012: using four approaches to characterize workers’ patterns of exposures": Table S3

| **Table S3. E-values for unmeasured confounding of the estimated risk ratios and 95% confidence intervals of work-related back injury, work-related illness, and fair-to-poor self-rated health from fully-adjusted city fixed effects models using *a priori* composite exposure, classification tree, and latent class approaches to defining exposure: NDWA-UIC CUED data, 14 cities, United States, 2011-2012 (N = 2,086).** | | | |
| --- | --- | --- | --- |
| **Variable** | **Risk ratio (95% confidence interval)** | **E-value for risk ratio** | **E-value for confidence interval limit closer to the null^a^** |
| ***Work-related back injury*** | | | |
| ***A priori* composite exposure** |  |  |  |
| Did no heavy lifting; no climbing to clean; did not work long hours with no break | Ref | Ref | Ref |
| Did heavy lifting or climbed to clean or worked long hours with no break | 3.2 (2.5 to 4.1) | 5.8 | 4.4 |
| Did heavy lifting and climbed to clean and worked long hours with no break | 6.5 (4.8 to 8.7) | 12.4 | 9.1 |
| **Classification tree** |  |  |  |
| Did no heavy lifting | Ref | Ref | Ref |
| Did heavy lifting | 3.0 (2.3 to 3.9) | 5.5 | 4.0 |
| Did heavy lifting and was verbally abused | 4.4 (3.6 to 5.3) | 8.2 | 6.6 |
| **Latent class analysis** |  |  |  |
| Low hazard domestic work | Ref | Ref | Ref |
| Demanding care work | 3.1 (2.0 to 4.9) | 5.6 | 3.3 |
| Strenuous cleaning work | 3.7 (2.6 to 5.2) | 6.8 | 4.6 |
| Hazardous domestic work | 6.6 (4.6 to 9.4) | 12.6 | 8.7 |
| ***Work-related illness*** | | | |
| ***A priori* composite exposure** |  |  |  |
| No contagious illness care; did not work long hours with no breaks | Ref | Ref | Ref |
| Did contagious illness care or worked long hours with no breaks | 1.9 (1.6 to 2.3) | 3.2 | 2.5 |
| Did contagious illness care and worked long hours with no breaks | 2.9 (2.4 to 3.6) | 5.3 | 4.1 |
| **Classification tree** |  |  |  |
| Did not work long hours with no breaks | Ref | Ref | Ref |
| Worked long hours with no breaks | 1.7 (1.4 to 2.0) | 2.8 | 2.2 |
| Worked long hours with no breaks and did contagious illness care | 2.6 (2.2 to 3.2) | 4.7 | 3.7 |
| **Latent class analysis** |  |  |  |
| Low hazard domestic work | Ref | Ref | Ref |
| Demanding care work | 2.2 (1.6 to 3.0) | 3.8 | 2.6 |
| Strenuous cleaning work | 2.1 (1.6 to 2.7) | 3.5 | 2.5 |
| Hazardous domestic work | 2.7 (2.0 to 3.5) | 4.8 | 3.5 |
| ***Fair-to-poor self-rated health*** | | | |
| ***A priori* composite exposure** |  |  |  |
| Did not work long hours with no breaks; did not work with toxic cleaning supplies; was not threatened | Ref | Ref | Ref |
| Worked long hours with no breaks or worked with toxic cleaning supplies or was threatened | 1.3 (1.1 to 1.5) | 1.9 | 1.5 |
| Worked long hours with no breaks and worked with toxic cleaning supplies and was threatened | 1.8 (1.4 to 2.3) | 2.9 | 2.0 |
| **Classification tree** |  |  |  |
| Did not climb to clean | Ref | Ref | Ref |
| Climbed to clean | 1.4 (1.2 to 1.7) | 2.2 | 1.8 |
| Climbed to clean and called insulting names or racial slurs | 1.9 (1.4 to 2.4) | 3.1 | 2.2 |
| **Latent class analysis** |  |  |  |
| Low hazard domestic work | Ref | Ref | Ref |
| Demanding care work | 1.0 (0.7 to 1.5) | 1.2 | 1.0 |
| Strenuous cleaning work | 1.5 (1.2 to 1.8) | 2.3 | 1.6 |
| Hazardous domestic work | 1.6 (1.3 to 2.0) | 2.6 | 1.8 |
| Abbreviations: NDWA-UIC CUED = National Domestic Workers Alliance and University of Illinois Chicago Center for Urban Economic Development. E-values represent the minimum strength of the association, on the risk ratio scale, that unmeasured confounder(s) would need to have with both the exposure and the outcome to fully explain away the estimated exposure-outcome association, conditional on all covariates included in the fully-adjusted health outcome models.  ^a^ “E-value for confidence interval limit closer to the null” refers to the E-value for the upper or lower bound of the confidence interval that is closer to the null value of, on the risk ratio scale, one. | | | |
