## Supplementary material for "Workplace hazards and health among informally employed domestic workers in 14 cities, United States, 2011-2012: using four approaches to characterize workers’ patterns of exposures": Figure S1

**Figure S1. Observed percent of domestic workers exposed to each single hazard (n=19 hazards), total and by selected individual, household, and occupational characteristics: NDWA-UIC CUED data, 14 cities, United States, 2011-2012 (N = 2,086).**

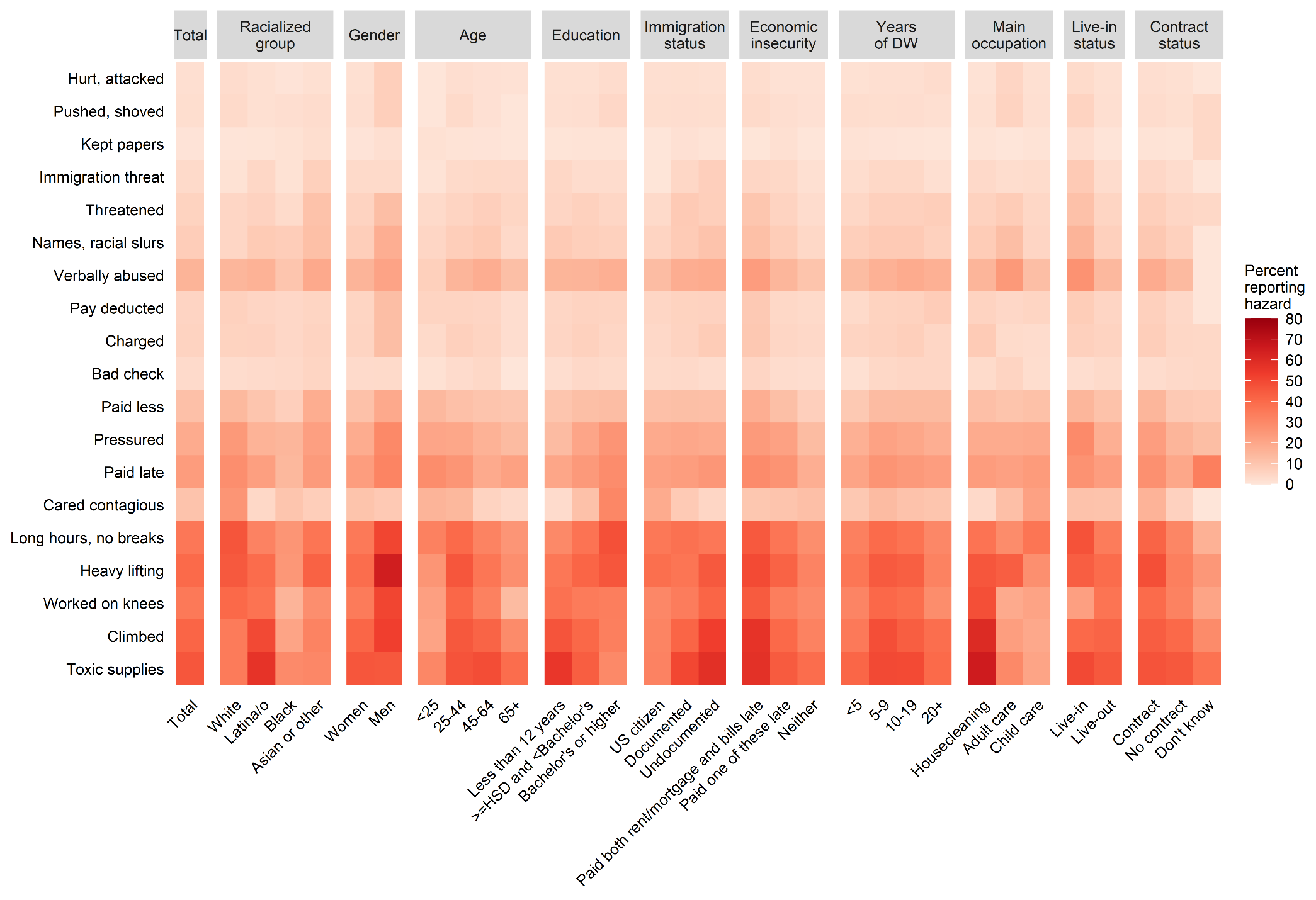

Abbreviations: DW = domestic work; HSD = high school diploma (or equivalent); NDWA-UIC CUED = National Domestic Workers Alliance and University of Illinois Chicago Center for Urban Economic Development; US = United States
