## Supplementary material for "Workplace hazards and health among informally employed domestic workers in 14 cities, United States, 2011-2012: using four approaches to characterize workers’ patterns of exposures": Figure S2

**Figure S2. Unpruned classification trees: NDWA-UIC CUED data, 14 cities, United States, 2011-2012 (N = 2,086).**

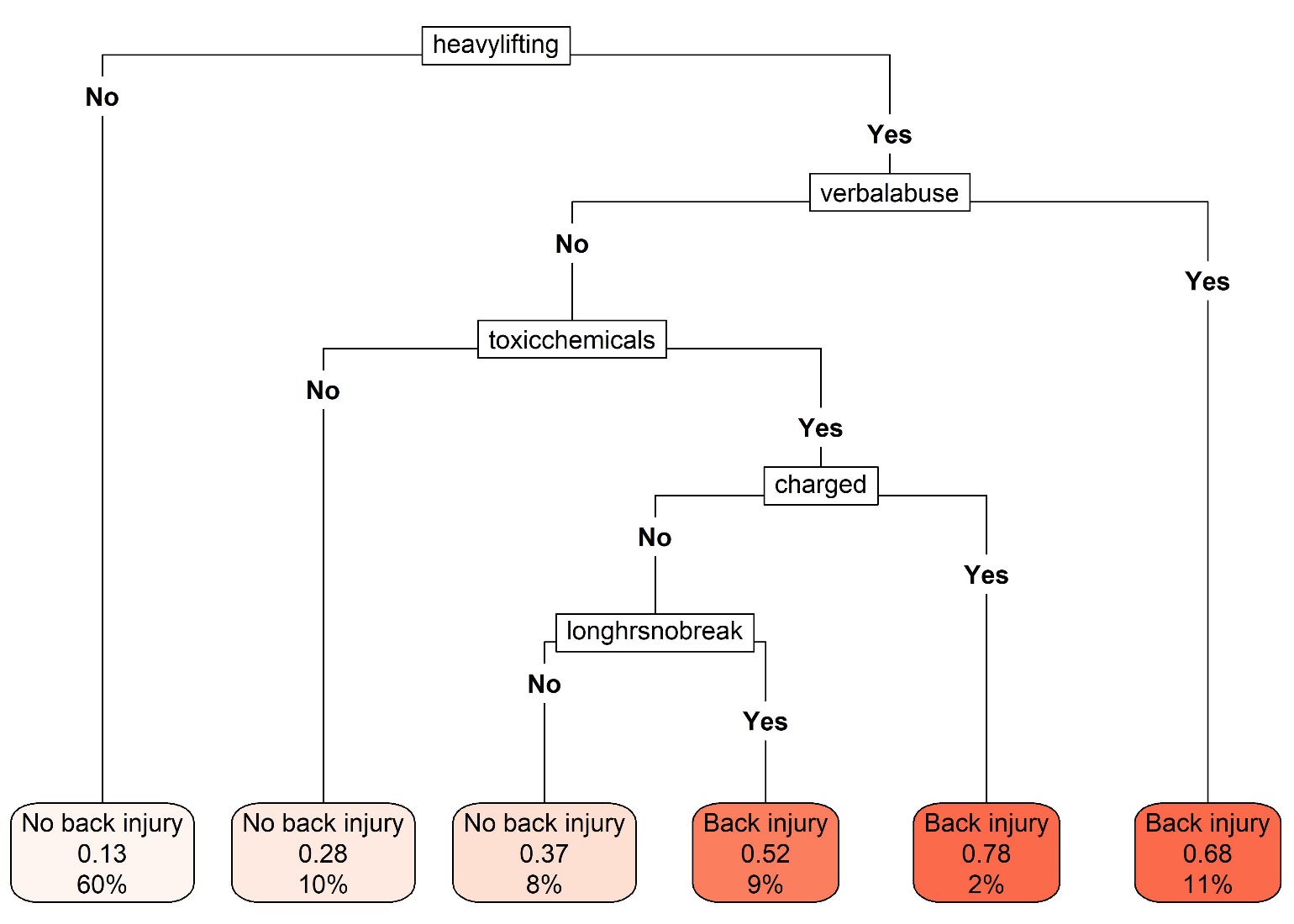
**A. Classification tree for work-related back injury**

| **B. Classification tree for work-related illness** | **C. Classification tree for fair-to-poor self-rated health** |
| --- | --- |
| 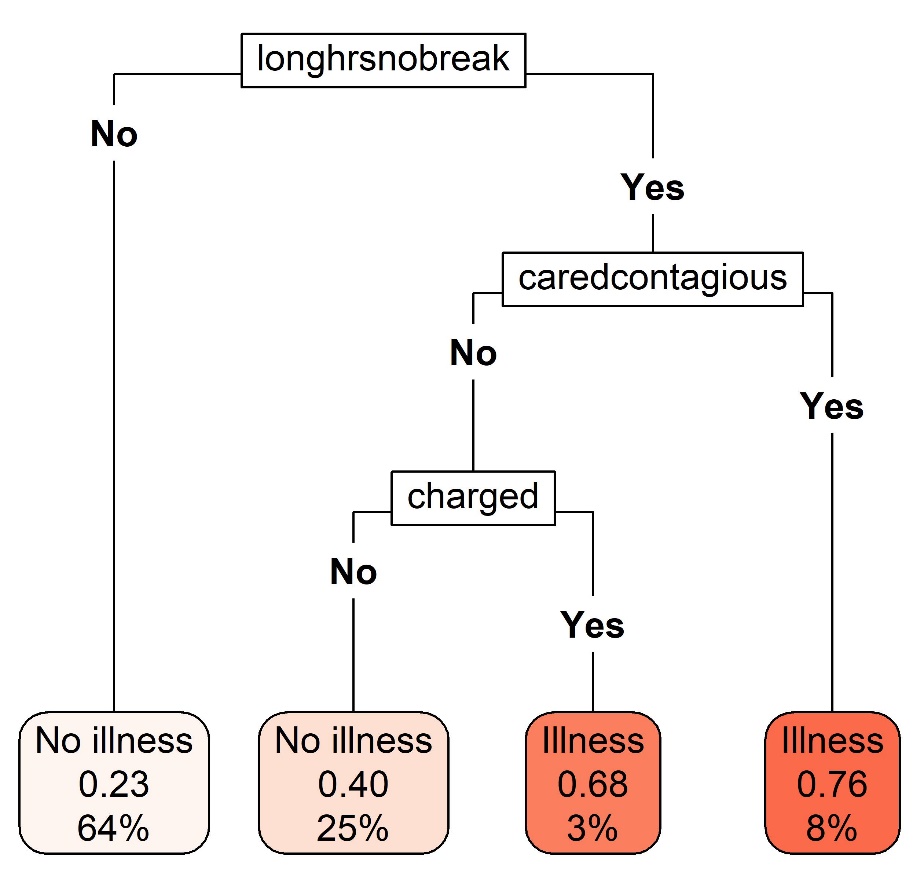 | 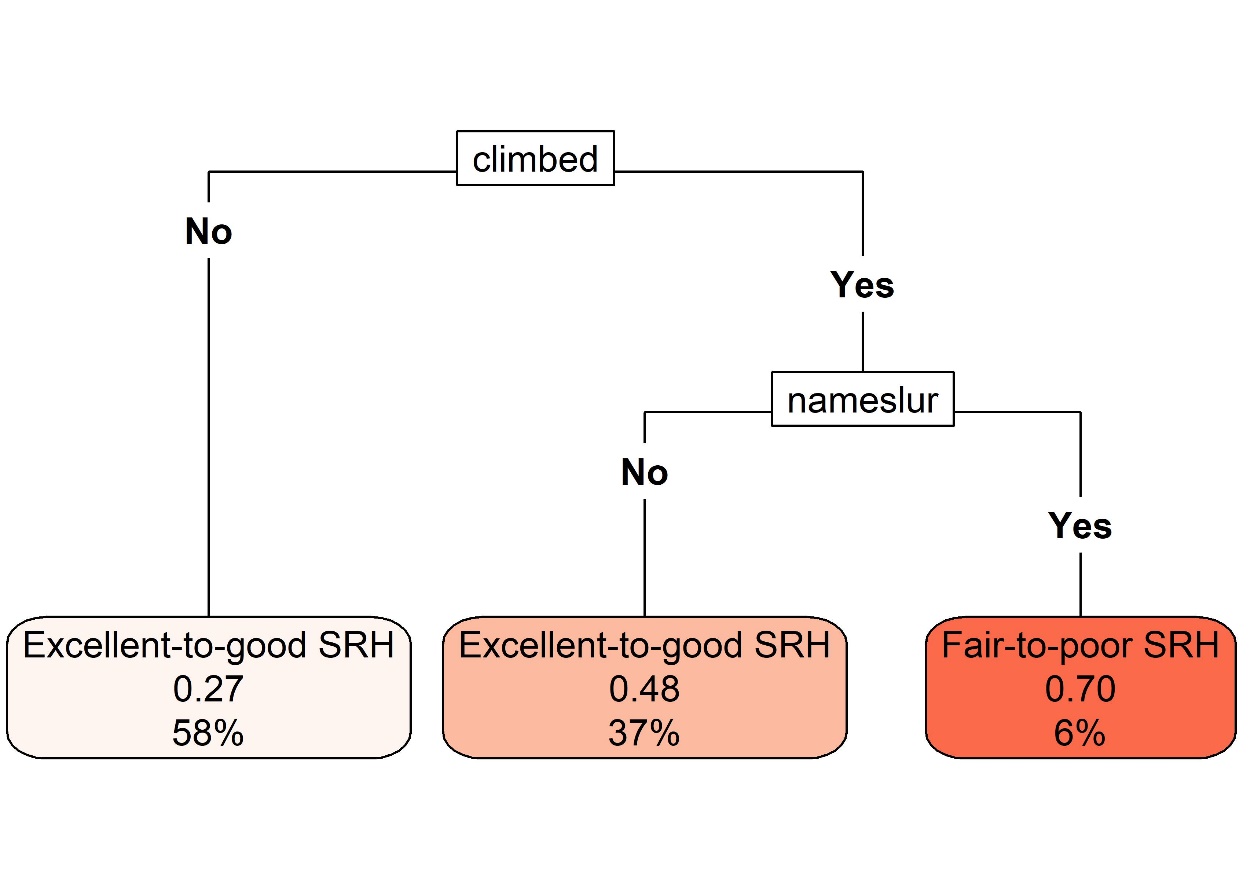 |
| Abbreviations: “caredcontagious” = Cared for someone with a contagious illness; “charged” = Charged for items that were broken or lost; “climbed” = Climbed to clean; “heavylifting” = Did heavy lifting; “longhrsnobreak” = Worked long hours with no breaks; “nameslur” = Called insulting names or racial slurs; NDWA-UIC CUED = National Domestic Workers Alliance and University of Illinois Chicago Center for Urban Economic Development; SRH = self-rated health; “toxicchemicals” = Worked with toxic cleaning supplies; “verbalabuse” = Verbally abused.  Figures show the unpruned classification trees fit using only the 19 hazard variables and no additional covariates. Each plot shows, in order of the elements of the figure from top to bottom: the name of the splitting variable (black textboxes), the splitting variable categories that determine how individuals are sorted based on that splitting variable (i.e., “No” exposure to a given hazard; “Yes” exposed to that hazard), and the final groups of individuals based on their responses to all splitting variables (shaded red textboxes). Within each final shaded red textbox is: a) the predicted health outcome (e.g., “No illness,” “Illness”), b) the predicted probability of experiencing the health outcome of interest (e.g., work-related back injury), and c) the percent of the total sample (n=2,086) assigned to that leaf of the tree. | |
