## Supplementary material for "Workplace hazards and health among informally employed domestic workers in 14 cities, United States, 2011-2012: using four approaches to characterize workers’ patterns of exposures": Figure S3

| **Figure S3. Pruned classification trees fit using covariates: NDWA-UIC CUED data, 14 cities, United States, 2011-2012 (N = 2,086).** | | |
| --- | --- | --- |
| 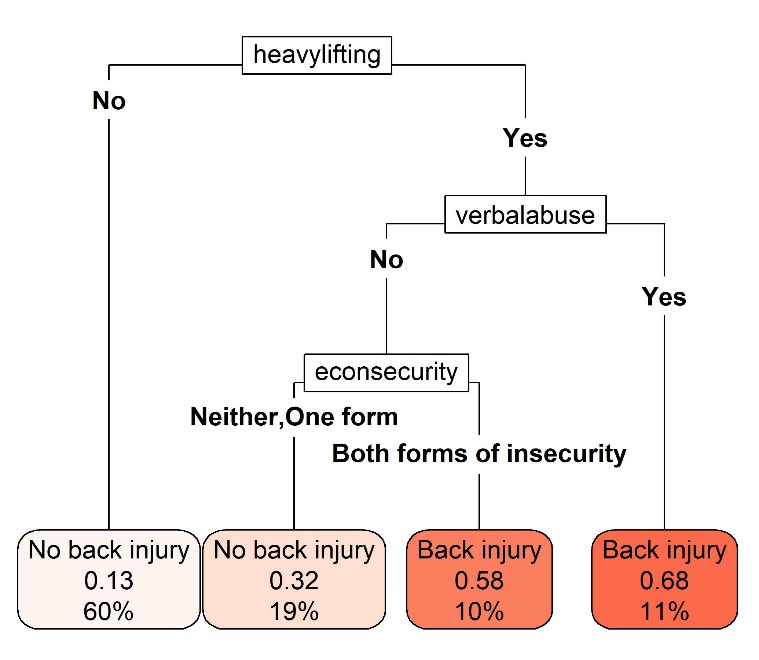**A. Classification tree for work-related back injury** | 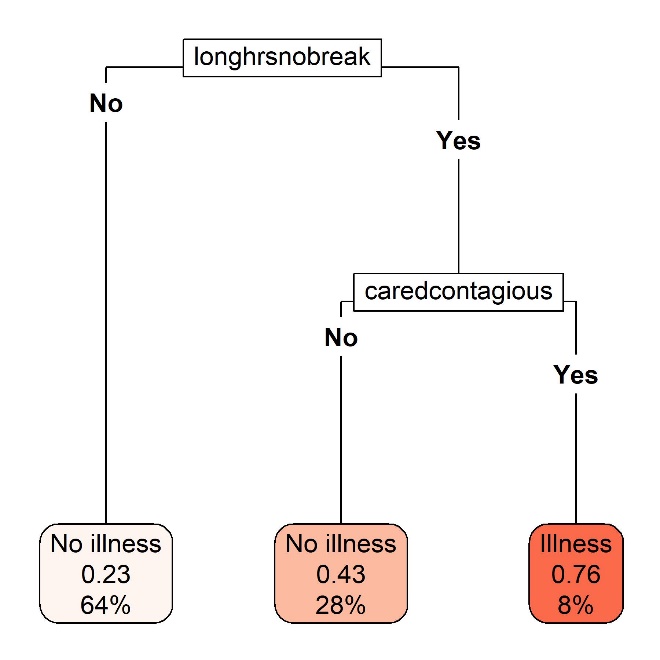**B. Classification tree for work-related illness** | 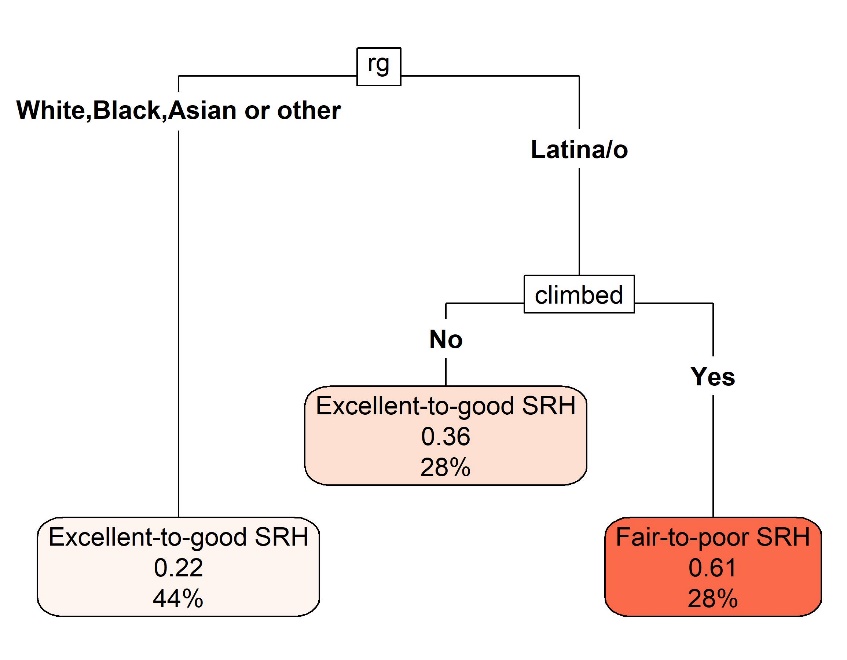**C. Classification tree for fair-to-poor self-rated health** |
| Abbreviations: “caredcontagious” = Cared for someone with a contagious illness; “climbed” = Climbed to clean; “econsecurity” = Household economic insecurity; “heavylifting” = Did heavy lifting; “longhrsnobreak” = Worked long hours with no breaks; NDWA-UIC CUED = National Domestic Workers Alliance and University of Illinois Chicago Center for Urban Economic Development; “rg” = Racialized group; SRH = Self-rated health; “verbalabuse” = Verbally abused.  Figures show the pruned classification trees fit using the 19 hazard variables and the following covariates: racialized group, formal educational attainment, citizenship and immigration status, age, main domestic work occupation, live-in status, and household economic insecurity. These are the same covariates included in the final latent class model. Each plot shows, in order of the elements of the figure from top to bottom: the name of the splitting variable (black textboxes), the splitting variable categories that determine how individuals are sorted based on that splitting variable (e.g., “No” exposure to a given hazard; “Yes” exposed to that hazard), and the final groups of individuals based on their responses to all splitting variables (shaded red textboxes). Within each final shaded red textbox is: a) the predicted health outcome (e.g., “No illness,” “Illness”), b) the predicted probability of experiencing the health outcome of interest (e.g., work-related back injury), and c) the percent of the total sample (n=2,086) assigned to that leaf of the tree. | | |
