## Supplementary material for "Workplace hazards and health among informally employed domestic workers in 14 cities, United States, 2011-2012: using four approaches to characterize workers’ patterns of exposures": Figure S4

**Figure S4. Estimated risk ratios and 95% confidence intervals of work-related back injury, work-related illness, and fair-to-poor self-rated health associated with each individual hazard (n=19 hazards) from fully-adjusted city fixed effects models: NDWA-UIC CUED data, 14 cities, United States, 2011-2012 (N = 2,086).**

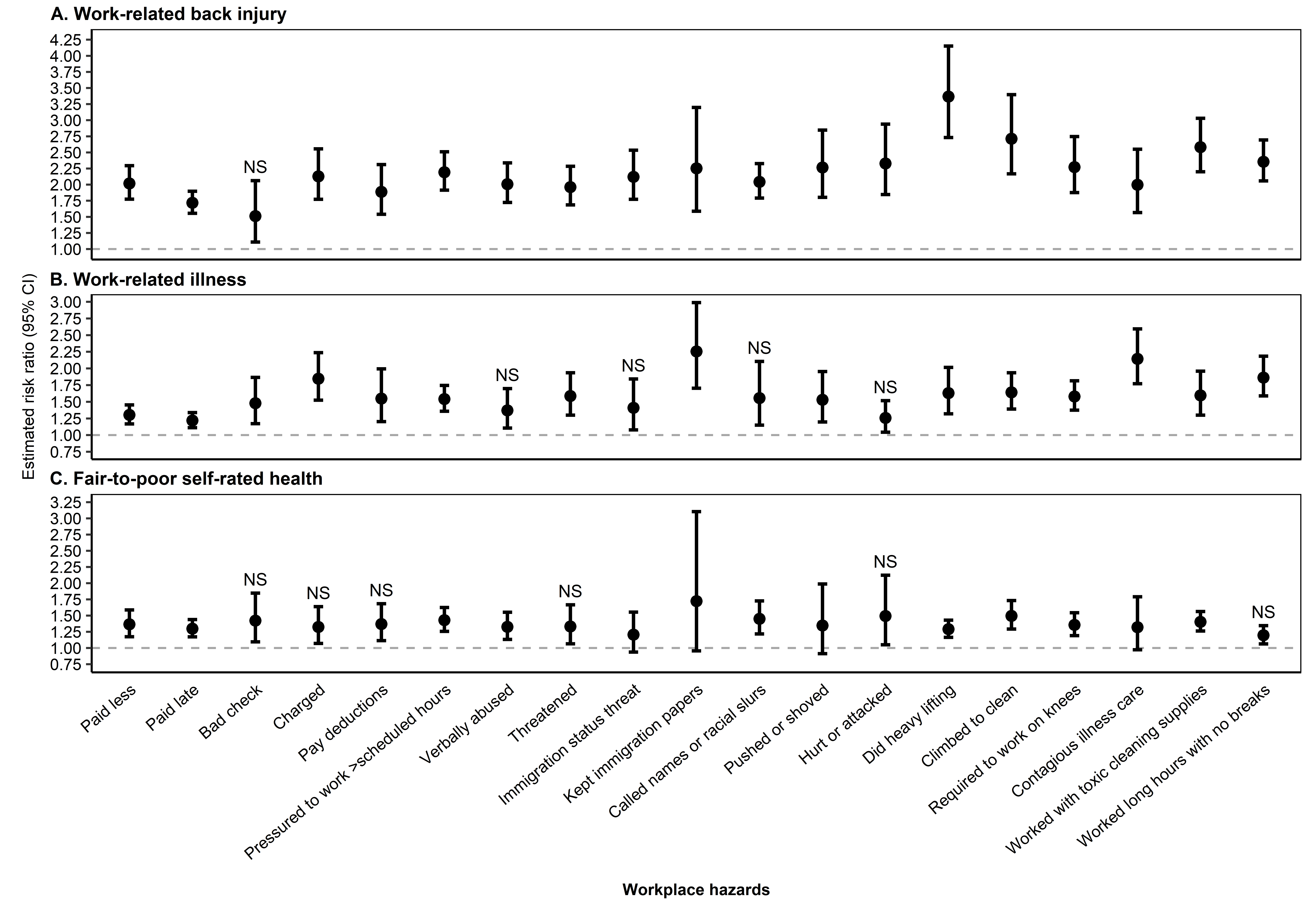

Abbreviations: CI = confidence interval; NDWA-UIC CUED = National Domestic Workers Alliance and University of Illinois Chicago Center for Urban Economic Development; NS = Not significant. A separate model was fit to obtain each of the point estimates and confidence intervals shown here. Each model included the indicated workplace hazard, city fixed effects, and was adjusted for the following selected covariates: racialized group, citizenship and immigration status, gender, age, formal educational attainment, main domestic work occupation, live-in status, years worked as a domestic worker in the United States, hours worked last week for one’s main domestic work employer, number of domestic work employers, had a contract with any of their domestic work employers in the last 12 months, number of people relying on the participant’s financial support, income earner status, and household economic insecurity. All confidence intervals are based on cluster robust standard errors, clustered at the city level. “NS” indicates estimates that are no longer statistically significant after adjusting for multiple testing of dependent hypotheses.
