## Supplementary material for "Workplace hazards and health among informally employed domestic workers in 14 cities, United States, 2011-2012: using four approaches to characterize workers’ patterns of exposures": Figure S5

**Figure S5. Estimated risk ratios and 95% confidence intervals—by which covariates are included in the models—of back injury, illness, and fair-to-poor self-rated health using *a priori* composite exposure, classification tree, and latent class methods: NDWA-UIC CUED data, 14 cities, United States, 2011-2012 (N = 2,086).**

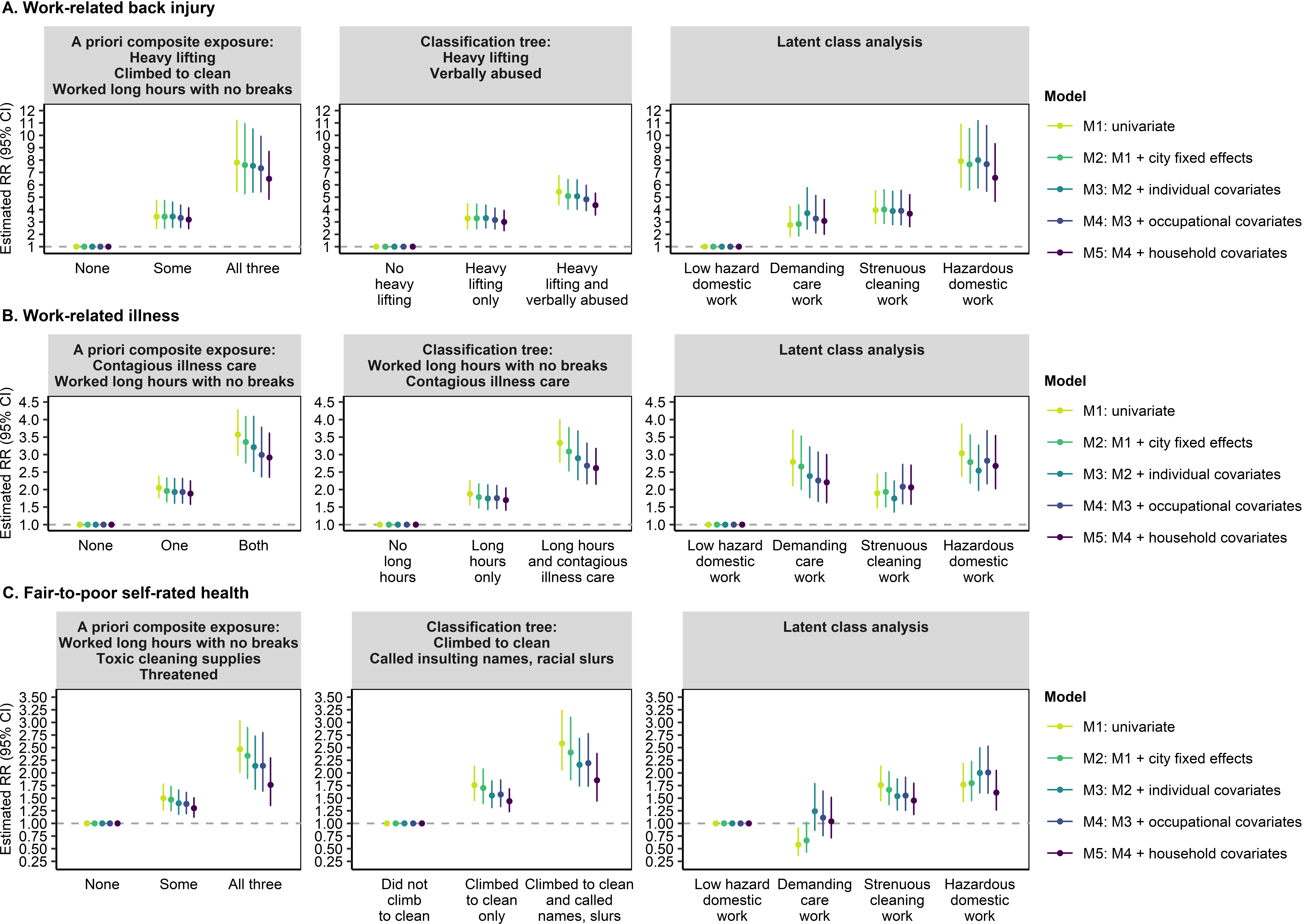

Abbreviations: CI = confidence interval; NDWA-UIC CUED = National Domestic Workers Alliance and University of Illinois Chicago Center for Urban Economic Development; RR = risk ratio. A separate model was fit to obtain each of the color-coded point estimates and confidence intervals shown here. M1 included only the exposure of interest. M2 additionally included the city fixed effects. M3 additionally included the individual covariates (racialized group, citizenship and immigration status, gender, age, and formal educational attainment). M4 additionally included the occupational covariates (main domestic work occupation, live-in status, years worked as a domestic worker in the United States, hours worked last week for one’s main domestic work employer, number of domestic work employers, had a contract with any of their domestic work employers in the last 12 months). M5 additionally included the household covariates (number of people relying on the participant’s financial support, income earner status, and household economic insecurity). All confidence intervals are based on cluster robust standard errors, clustered at the city level.
