## Supplementary material for "Workplace hazards and health among informally employed domestic workers in 14 cities, United States, 2011-2012: using four approaches to characterize workers’ patterns of exposures": Figure S6

**Figure S6. Proportion of domestic workers with selected characteristics in each exposure category, as defined by the four approaches and in relation to each or all of the health outcomes: National Domestic Workers Alliance and University of Illinois Chicago Center for Urban Economic Development data, 14 cities, United States, 2011-2012 (N = 2,086).**

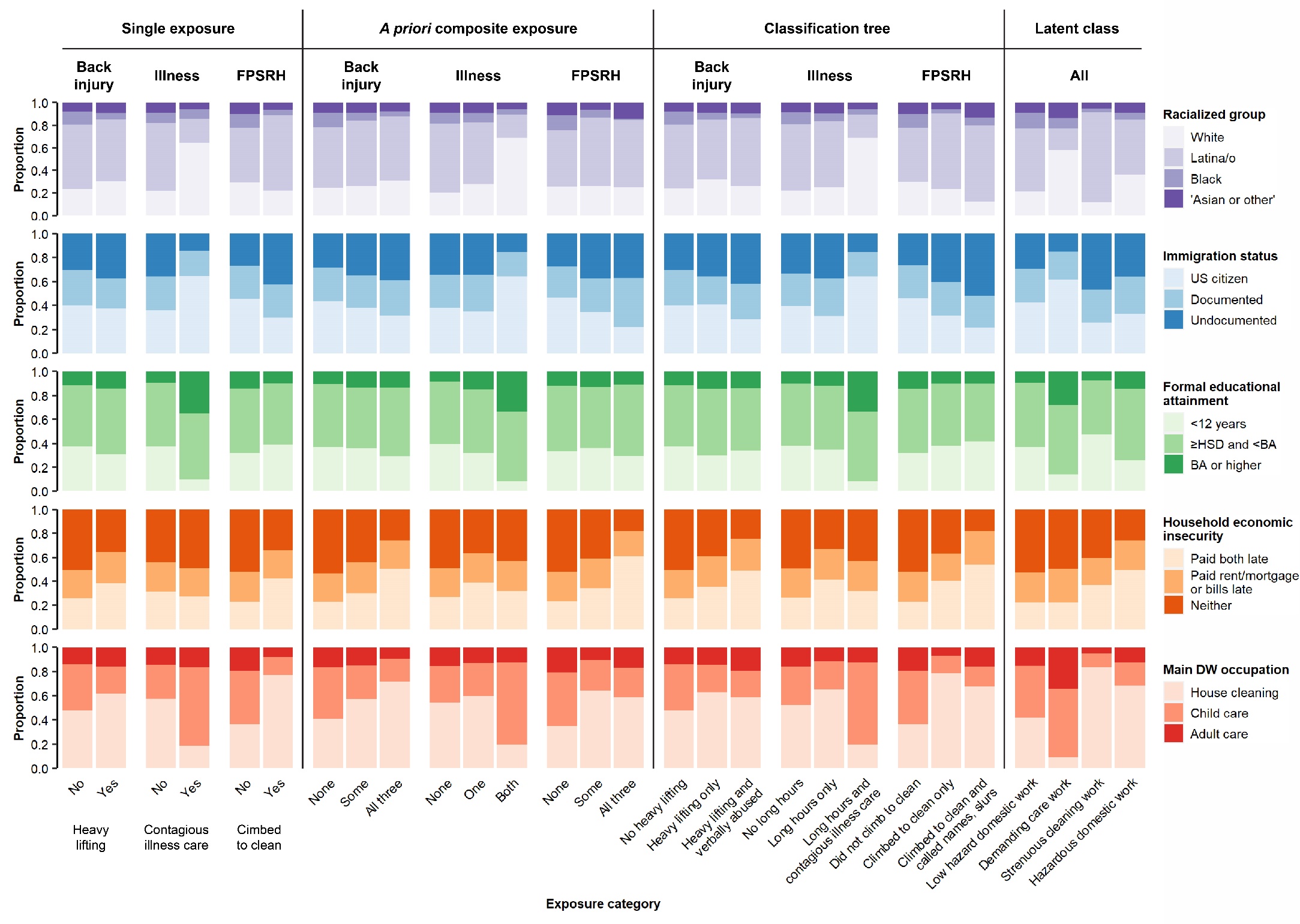

Abbreviations: BA = Bachelor’s degree; DW = domestic work; FPSRH = fair-to-poor self-rated health; HSD = high school diploma (or equivalent); US = United States.

*A priori* composite exposure categories defined in relation to back injury are as follows: None = No heavy lifting AND did not climb to clean AND did not work long hours with no breaks; Some = Heavy lifting OR climbed to clean OR worked long hours with no breaks; All = Heavy lifting AND climbed to clean AND worked long hours with no breaks.

*A priori* composite exposure categories defined in relation to illness are as follows: None = No contagious illness care AND did not work long hours with no breaks; Some = Contagious illness care OR worked long hours with no breaks; All = Contagious illness care AND worked long hours with no breaks.

*A priori* composite exposure categories defined in relation to FPSRH are as follows: None = Did not work long hours with no breaks AND did not work with toxic cleaning supplies AND was not threatened; Some = Worked long hours with no breaks OR worked with toxic cleaning supplies OR was threatened; All = Worked long hours with no breaks AND worked with toxic cleaning supplies AND was threatened.
